# Lipidomics for diagnosis and prognosis of pulmonary hypertension

**DOI:** 10.1101/2023.05.17.23289772

**Authors:** Natalie Bordag, Bence Miklos Nagy, Elmar Zügner, Helga Ludwig, Vasile Foris, Chandran Nagaraj, Valentina Biasin, Ulrich Bodenhofer, Christoph Magnes, Bradley A. Maron, Silvia Ulrich, Tobias J. Lange, Konrad Hötzenecker, Thomas Pieber, Horst Olschewski, Andrea Olschewski

## Abstract

**Background:** Pulmonary hypertension (PH) poses a significant health threat with high morbidity and mortality, necessitating improved diagnostic tools for enhanced management. Current biomarkers for PH lack functionality and comprehensive diagnostic and prognostic capabilities. Therefore, there is a critical need to develop biomarkers that address these gaps in PH diagnostics and prognosis.

**Methods:** To address this need, we employed a comprehensive metabolomics analysis in 233 blood based samples coupled with machine learning analysis. For functional insights, human pulmonary arteries (PA) of idiopathic pulmonary arterial hypertension (PAH) lungs were investigated and the effect of extrinsic FFAs on human PA endothelial and smooth muscle cells was tested *in vitro*.

**Results:** PA of idiopathic PAH lungs showed lipid accumulation and altered expression of lipid homeostasis-related genes. In PA smooth muscle cells, extrinsic FFAs caused excessive proliferation and endothelial barrier dysfunction in PA endothelial cells, both hallmarks of PAH.

In the training cohort of 74 PH patients, 30 disease controls without PH, and 65 healthy controls, diagnostic and prognostic markers were identified and subsequently validated in an independent cohort. Exploratory analysis showed a highly impacted metabolome in PH patients and machine learning confirmed a high diagnostic potential. Fully explainable specific free fatty acid (FFA)/lipid-ratios were derived, providing exceptional diagnostic accuracy with an area under the curve (AUC) of 0.89 in the training and 0.90 in the validation cohort, outperforming machine learning results. These ratios were also prognostic and complemented established clinical prognostic PAH scores (FPHR4p and COMPERA2.0), significantly increasing their hazard ratios (HR) from 2.5 and 3.4 to 4.2 and 6.1, respectively.

**Conclusion:** In conclusion, our research confirms the significance of lipidomic alterations in PH, introducing innovative diagnostic and prognostic biomarkers. These findings may have the potential to reshape PH management strategies.

## Introduction

Pulmonary hypertension (PH) affects 1% of the world’s population^1,2^ and thus represents a significant global health problem. Even mild PH is a strong negative prognosticator in patients with left heart disease^3^, lung disease^4^, and pulmonary arterial hypertension (PAH)^5,6^.

In recent decades, global research efforts have led to targeted therapies for the rare pulmonary vascular diseases PAH and chronic thromboembolic PH (CTEPH). Despite this, the estimated five-year survival rate for newly diagnosed PAH patients has remained at only 61%^7,8^.

Diagnosis of PH is challenging because measurement of pulmonary arterial pressure (PAP) requires right heart catheterization (RHC), whereas non-invasive methods provide only reliable estimates of PAP^1^ or are not widely available^9^. Natriuretic peptide levels (BNP or NT-proBNP) are the only recommended biomarkers for PH, but they are not specific for pulmonary hypertension. Therefore, the development of new diagnostic tools for the detection of PH, risk stratification, and epidemiological studies remains an important issue^2^.

Fibroproliferative remodelling of distal pulmonary arterioles drives elevation in pulmonary vascular resistance and pulmonary arterial pressure. This is associated with unique metabolic changes as detected from the circulation^10–15^ as a reflection of the profound changes in the cells and matrix of the right ventricle and pulmonary vessel walls^16^. Our hypothesis was that PH patients may present with a characteristic metabolic profile that allows for detection of PH and risk stratification^1^. We identified lipidomic changes in the small pulmonary arteries (PA) of IPAH patients and a unique lipidomic profile in a diverse PH cohort, allowing identification of PH among healthy and diseased controls, which was confirmed in an independent validation cohort. In addition, we show that simple markers of this lipidomic profile are significantly associated with survival and improve the accuracy of two established prognostic PAH scores.

## Results

### Clinical and cardiopulmonary hemodynamic characteristics for the study cohorts

Our study included three classes of subjects: PH patients, healthy control subjects (HC) and lung disease controls (DC) without PH, all of whom underwent blood-based high resolution mass spectrometry (HRMS)-based metabolomics (see Fig. 1). PH patients had a mean PAP (mPAP) ≥ 25 mmHg and were either 1) PAH, 2) PH due to left heart disease (LV), 3) PH due to lung disease, or 4) CTEPH. All DC had mPAP < 25 mmHg with either metabolic syndrome, chronic obstructive pulmonary disease (COPD) or interstitial lung disease (ILD). The distributions in sex and body mass index (BMI) between PH and HC/DC were similar (see Fig. 1C). Age was in the same range, although controls tended to be younger. Metabolites were not significantly correlated with age (see Fig. S3).

**Fig. 1.**
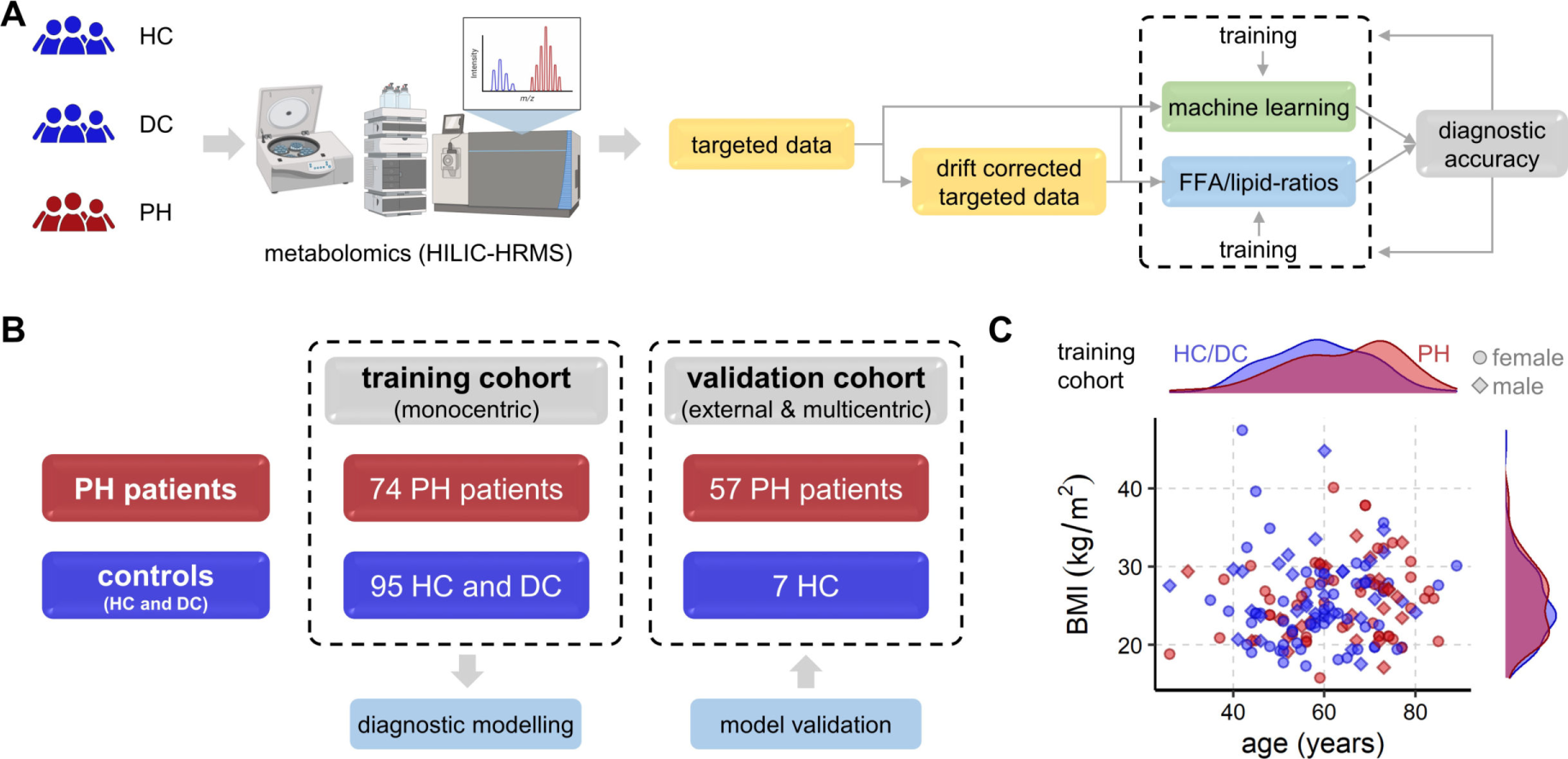
Study overview for all cohorts. **(A)** Schematic workflow of metabolomics measurement and computational analysis. Created with BioRender.com. **(B)** Schematic overview of group distribution of all included patients (n = 233) in training and validation cohorts. **(C)** Scatter plot of BMI vs. age (shape of symbols by sex) in the training cohort with distribution histograms per PH and per HC/DC showing comparable distributions of sex and BMI in PH and HC/DC, avoiding potential confounders by design.

The training cohort consisted of PH patients, DC and HC, all sampled in Graz, Austria. The external validation cohort contained PH patients and HC from Zürich, Switzerland and Regensburg, Germany. Patient characteristics are summarized in Table 1.

**Table 1:** Patients’ characteristics (medians ± 95% confidence interval)

|  | Training (n=169) |  |  | Validation (n=64) |  |
| --- | --- | --- | --- | --- | --- |
|  | HC (n=65) | DC (n=30) | PH (n=74) | HC (n=7) | PH (n=57) |
| Age, years | 58.0±2.8 | 59.5±4.7 | 66.5±3.0 | 42.0±13.8 | 57.0±3.7 |
| Female:male (ratio) | 44:21 (2.1:1) | 20:10 (2.0:1) | 50:24 (2.1:1) | 5:2 (2.5:1) | 43:14 (3.1:1) |
| BMI (kg/m<sup>2</sup>) | 24.1±0.9 | 26.2±3.0 | 25.9±1.1 | 21.0±1.7 | 26.3±1.4 |
| Diagnosis (since years) | - | 8±1.8 | 1.0±1.2 | - | 5.5±1.7 |
| *Pulmonary hemodynamics from RHC* |  |  |  |  |  |
| mPAP (mmHg)<br>mean pulmonary arterial pressure | - | - | 42.0±2.5 | - | 50.0±3.9 |
| PAWP (mmHg)<br>pulmonary arterial wedge pressure | - | - | 10.0±1.3 | - | 9.0±0.9 |
| CO (L/min)<br>cardiac output | - | - | 4.3±0.4 | - | 4.2±0.4 |
| CI (L/min/m<sup>2</sup>)<br>cardiac index | - | - | 2.6±0.2 | - | 2.4±0.2 |
| PVR (WU)<br>pulmonary vascular resistance | - | - | 7.1±1.0 | - | 9.1±1.7 |
| RAP (mmHg)<br>right atrial pressure | - | - | 6.0±1.2 | - | 7.5±1.1 |
| *Clinical data* |  |  |  |  |  |
| 6MWD (m)<br>6-min walk distance | - | 454±41 | 330±34 | - | 390±31 |
| WHO FC<br>world health organisation functional class | - | 2.0±0.2 | 3.0±0.2 | - | 2.0±0.2 |
| FEV1 (% predicted)<br>forced expiratory volume 1 s | - | 51.5±9.9 | 75.0±5.6 | - | 82.2±3.0 |
| FVC (% predicted)<br>forced vital capacity | - | 63.8±6.7 | 82.8±5.5 | - | 91.4±4.0 |
| FEV1/FVC (% predicted) | - | 56.3±8.5 | 74.4±3.0 | - | 75.7±2.1 |
| TLC (% predicted)<br>total lung capacity | - | 103.0±10.8 | 94.0±3.9 | - | 96.5±3.5 |
| DLCO cSB (% predicted)<br>single-breath CO diffusing capacity,<br>haemoglobin corrected | - | 49.6±7.8 | 63.8±6.0 | - | 53.8±3.9 |
| DLCO cVA (% predicted)<br>diffusing capacity for CO alveolar<br>volume, haemoglobin corrected | - | 70.0±7.9 | 69.6±6.1 | - | 62.0±3.7 |
| RDW (%)<br>red cell distribution width | - | 14.0±0.7 | 15.4±0.7 | - | 14.7±0.6 |
| NT-proBNP (pg/mL) | - | 98±39 | 869±806 | - | 751±475 |
| Uric acid (mg/dL) | - | 5.0±0.5 | 6.0±0.6 | - | 6.6±0.7 |
| Creatinine (mg/dL) | - | 0.80±0.09 | 1.02±0.12 | - | 1.00±0.23 |
| Bilirubin total (mg/dL) | - | 0.40±0.13 | 0.64±0.13 | - | 0.65±0.16 |

### Identification of a characteristic lipidomics profile in pulmonary hypertension

The metabolome of patients was assessed with untargeted hydrophilic interaction liquid chromatography (HILIC)-HRMS from serum, EDTA, and heparin plasma samples in four measurement runs. As is typical with mass spectrometry (MS) based metabolomics methods, notable batch effects occurred between runs as well as drift within the longer runs (see Fig. S2). Drift correction was successfully performed (see Fig. S2) using quality control (QC) injections, which are a mix of equal sample volumes repeatedly measured to monitor instrument stability.

In total, 164 known metabolites were of consistent analytical quality suitable for multivariate and univariate exploratory analysis. Global metabolic changes were first examined using the unsupervised multivariate independent principal component analysis (iPCA). The metabolomes of the PH patients differed from the control groups (HC and DC), which was visible as a clear group separation in the iPCA scores plot along the first component (x-axis, Fig. 2A). The observed metabolic difference was strongly driven by an increase in specific free fatty acids (FFA) in PH patients (Fig. 2B). The machine learning method orthogonal projections to latent structures discriminant analysis (OPLS-DA) confirmed that the observed global metabolic differences between PH and HC/DC were significant (p < 0.001, cross-validation and 1000 random permutations, Fig. 2C).

**Fig. 2.**
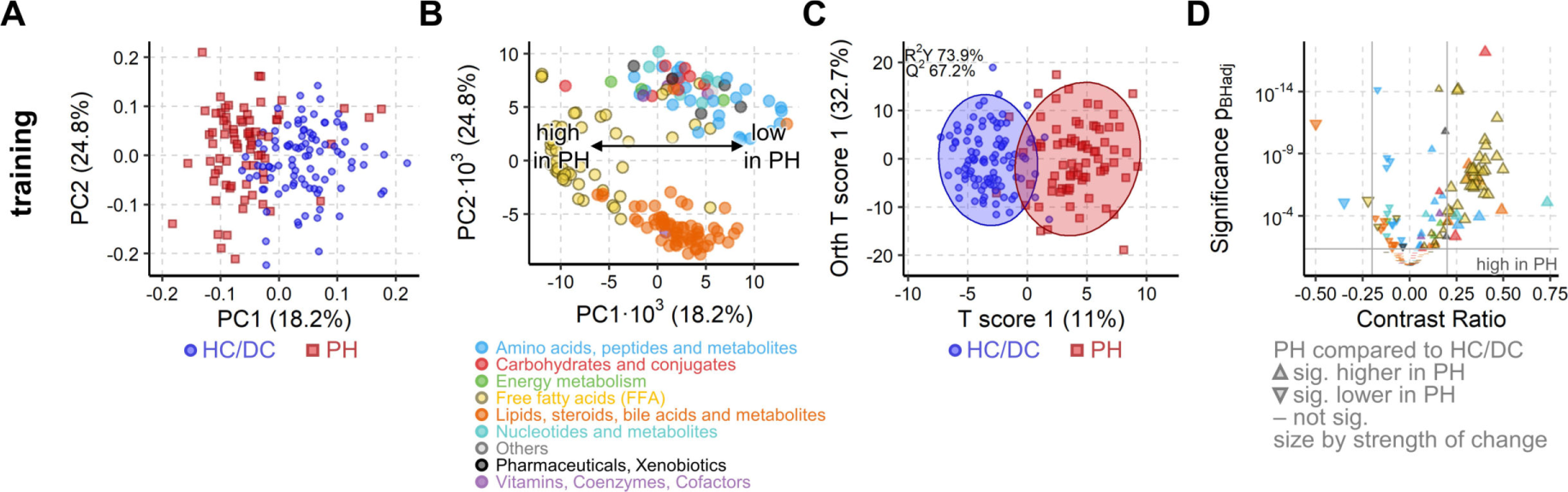
PH is associated with a strong metabolic shift in the training cohort. **(A)** iPCA scores plot representing the metabolic profile of each subject as a dot. The proximity of the dots indicates the similarity of the subjects’ metabolomes. Clear group separation by PH is visible along the first component. **(B)** Loadings plot corresponding to scores plot in (A). Each dot represents the contribution of the metabolite to the group separation observed in the scores plot. FFAs (yellow circles) strongly drive the group separation and are increased in PH patients. **(C)** OPLS-DA maximizes the group difference from PH to HC/DC and the resulting scores plot represents, as in A, the metabolome of each subject. Similarly, proximity indicates similarity and ellipses mark the 95% confidence interval of the groups. The difference between the metabolome of PH and HC/DC was significant (Q^2^ > 50%, p< 0.001 from 1000 random permutation). **(D**) Volcano plot of univariate analysis showing significant (p_BH_ < 0.05, grey horizontal line) and strong (absolute contrast ratio > 0.25, grey vertical lines) increase in FFAs (yellow triangles). For all methods A-D, 164 known metabolites from the training cohort samples (n = 169) were used (drift corrected, *log_10_*-transformed data). Colors as in B.

The univariate statistical analysis confirmed that FFAs were strongly and significantly increased in PH as compared with HC/DC (Fig. 2D). The metabolites from routine clinical chemistry, e.g. uric acid, were strongly correlated with their respective HILIC-HRMS metabolites (Fig. S3). Single FFAs and lipids were not strongly correlated with any clinical parameter, suggesting that the detected FFA changes may be independent from conventional clinical assessment.

### FFA/lipid-ratios diagnose pulmonary hypertension

The potential of using our metabolites to predict PH was first investigated with the machine learning method random forest (RF)^17,18^ and extreme gradient boosting (XGBoost)^19^. From the 164 metabolites, 11 were excluded from biomarker analysis because of low signal intensities and high noise (see Supplementary Data 1). RF and XGBoost both achieved an area under the curve (AUC) of 0.82 in the receiver operator curve (ROC) analysis in the validation set (Fig. 3A). The drift correction used here allowed a joint exploratory statistical analysis, but drift correction is impossible in future routine clinical diagnostics. Therefore, the diagnostic and prognostic performance was also tested without drift correction. RF and XGBoost performance were almost identical without drift correction, indicating that their nonlinear algorithms exhibit intrinsic drift-handling capabilities (Fig. 3A).

**Fig. 3.**
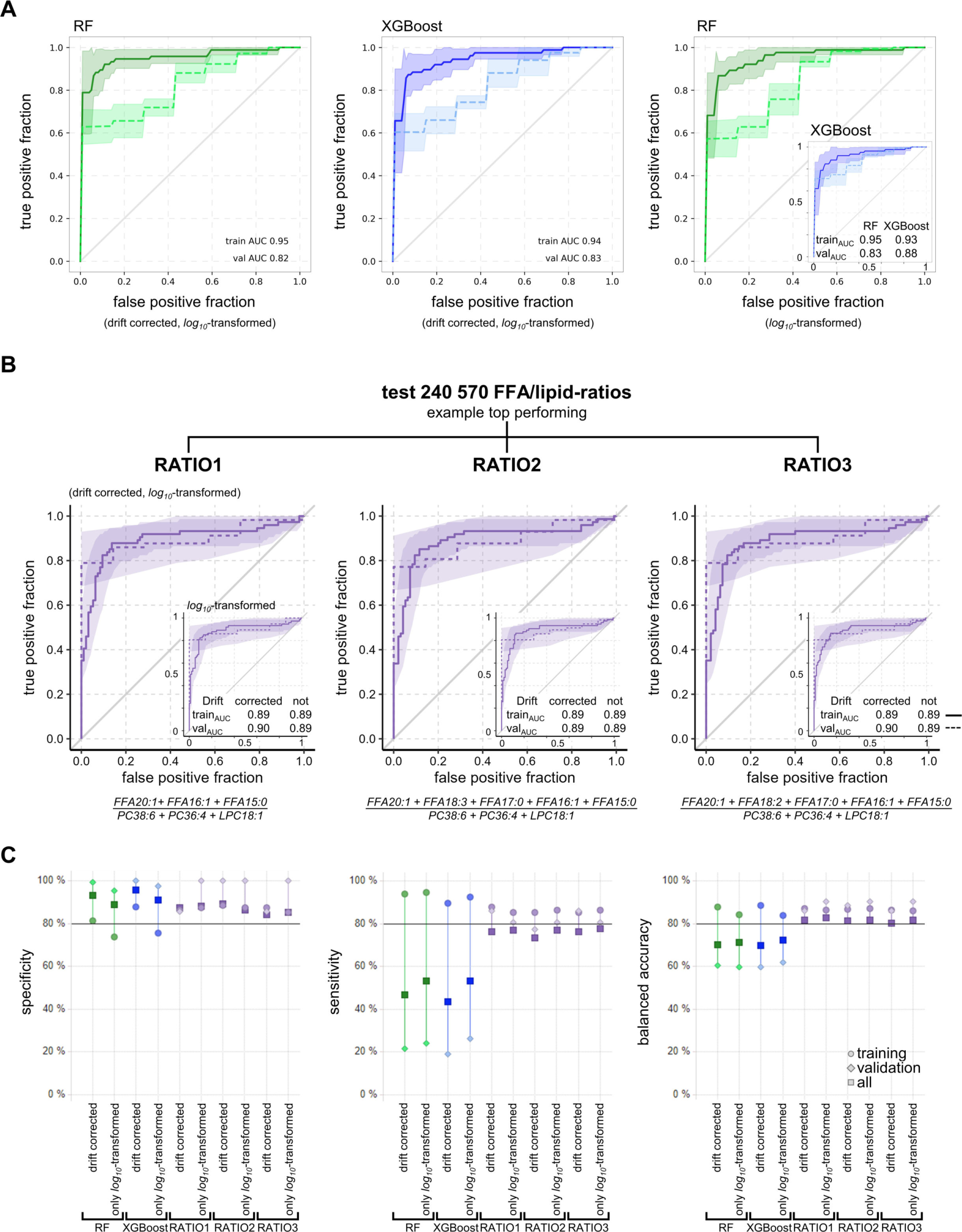
Diagnostic accuracy for PH in training and validation cohorts. **(A)** ROC plots of RF (green) and XGBoost (blue) trained with data from training cohort predicting class in validation cohort with either drift corrected, *log_10_*-transformed data (left, middle) or non-drift corrected *log_10_*-transformed data (right) based on 153 metabolites. **(B)** The ROC plots of the three best FFA/lipid-ratios for training and validation cohort with either drift corrected, *log_10_*-transformed data or *log_10_*-transformed data with no drift correction (insets). **(A-B)** Training cohort n = 169 (solid line), validation cohort n = 64 (dashed line), ribbons mark 95% confidence intervals. **(C)** Plot of model performance metrics sensitivity, specificity and balanced accuracy for RF, XGBoost, and the three best FFA/lipid-ratios when based on either training (circles) or validation (diamonds) cohorts only or all available data (squares). The performance of RF and XGBoost was comparable for all three metabolite subsets with and without drift correction. The performance of the three best FFA/lipid-ratios was consistently more stable and balanced than of RF or XGBoost.

Other important model performance parameters such as specificity, sensitivity, and balanced accuracy were comparable for RF and XGBoost irrespective of drift correction. For RF and XGBoost, the average balanced accuracy was 72.2% and 72.7%, respectively, the specificity was 88.6% and 91.2%, respectively, and the sensitivity was 55.8% and 54.1%, respectively (Fig. 3C). The validation cohort had only seven HC and no DC with a slightly different age and BMI distribution than the training cohorts. To overcome this limitation, we tested an artificial data split into 70% training and 30% validation sets with balanced distributions in age, BMI, sex and disease class (PH/DC/HC). The performance of RF and XGBoost remained similar to the original split by center (Fig. S4).

Despite their diagnostic potential, both machine learning approaches are not suitable for routine clinical diagnostics because they are labour intensive and not fully explainable. In addition, both approaches showed notable decreases in AUC, sensitivity, specificity and balanced accuracy from the training to the validation cohort. Therefore, we tested whether ratios formed from lipophilic metabolites with strong effects of PH versus HC/DC, normalized to metabolites with no effects of PH vs. HC/DC, could replace machine learning approaches to create an explainable, easy-to-measure marker. In PH, many FFAs were strongly increased while many complex lipids were unchanged (Fig. 2D), offering the option to achieve markers of PH that are easy to measure. Thus, characteristic FFAs were selected into the numerator, based on their analytical performance in PH versus HC/DC and lipids with good analytical performance and non-significance in PH vs. HC/DC were chosen for the denominator.

For the nominator, 11 FFA were considered: FFA C15:0 (pentadecylic acid), FFA C16:2 (palmitolinoleic acid), FFA C16:1 (palmitoleic acid), FFA C17:1 (heptadecenoic acid), FFA C17:0 (margaric acid), FFA C18:3 (α-linolenic acid, ALA, or γ-linolenic acid, GLA), FFA C18:2 (linoleic acid, LA), FFA C18:1 (oleic acid), FFA C19:1 (nonadecenoic acid), FFA C20:5 (eicosapentanoic acid, EPA) and FFA C20:1 (eicosenoic acid). Eight lipids, the best two from four common classes, were considered for the denominator: lysophosphatidylcholine (LPC) 18:2, LPC 18:1, phosphatidylcholine (PC 36:4, PC 38:6), sphingomyelin (SM 34:2, SM 36:2), lysophosphatidylethanolamine (LPE) 16:0, and LPE 18:1. To stabilize the ratios, we also tested sums of up to six FFAs in the numerator and up to four lipids in the denominator, limiting the maximum number of individual metabolites in a given ratio to ten.

In total, about a quarter of a million FFA/lipid-ratios were evaluated for their diagnostic performance in ROC analysis. Most FFA/lipid-ratios achieved moderate to high performance with AUCs above 0.8 (Fig. S5A-I,K,L) irrespective of drift correction or split. There may be a concern that combining multiple metabolites in a ratio increases technical error and impairs reproducibility. However, in our cohort, the calculated technical variability according to the laws of error propagation was <5% for most FFA/lipid-ratios and <10% for all others (Fig. S5J).

For simplicity, the top three ratios were selected, based on their AUC, sensitivity, and specificity, and named RATIO1, RATIO2, and RATIO3 (Fig. S5M). These ratios had six metabolites in common: numerator (FFA C20:1 + FFA C16:1 + FFA C15:0); denominator (PC 38:4 + PC 36:4 + LPC 18:1). Compared to RATIO 1, the only difference of RATIO 2 and 3 was the addition of two more FFA to the numerator sum (FFA 18:3 + FFA 17:0 and FFA 18:2 + FFA 17:0, Fig. 3B). The ROC curves of RATIO1-3 overlapped well between training and validation cohort, irrespective of drift correction (Fig. 3B). All other important performance parameters were also very similar between RATIO1-3 (Fig. 3C). As a general result, top ratios outperformed the machine learning models, irrespective of drift correction or data split, especially in terms of sensitivity. The top three ratios’ average balanced accuracy was 85.5%, with 89.7% specificity and 81.4% sensitivity (Fig. 3C).

FFA/lipid-ratios massively reduced technical complexity compared to broad metabolomics runs while achieving better diagnostic performance than machine learning approaches. The diagnostic performance of the FFA/lipid-ratios was independent of drift correction, making them suitable for stand-alone measurements. Accordingly, we assume that FFA/lipid-ratios are suitable for future routine applications.

### Specific FFA/lipid-ratios predict survival

The prognostic value of RATIO1 was compared with well-established prognostic PAH scores, FPHR4P^20^ (based on WHO FC, 6MWD, RAP, and CI) and COMPERA2.0^1,21^ (based on WHO FC, 6MWD, and NT-pro-BNP). Survival and hazard ratios (HR) were investigated for all our patients with heart or lung disease and PH. For direct comparability of Kaplan-Meier survival curves and HRs, all numeric values were categorized into low or high risk according to their optimal cut-off points (e.g. 57 years for age).

We analysed survival times since enrolment (= baseline) which were available for 129 PH and 21 DC (13 COPD, 8 ILD) patients. The COMPERA2.0 score was available for 122 PH and 11 DC patients, and FPHR4p for 97 patients (93 PH and 4 DC). Survival times and both scores were available for 91 PH patients. As expected, FPHR4p and COMPERA2.0 scores were significantly associated with survival time (Fig. 4). RATIO1 was also significantly associated with survival (Fig. 4) with a similar HR as COMPERA2.0 scores. When RATIO1 was combined with each of the scores (RATIO1 and score equally weighted), the prognostic value of the respective score improved notably (Fig. 4). This indicates that our simple metabolomic marker provided independent prognostic information and would complement established prognostic scores.

**Fig. 4.**
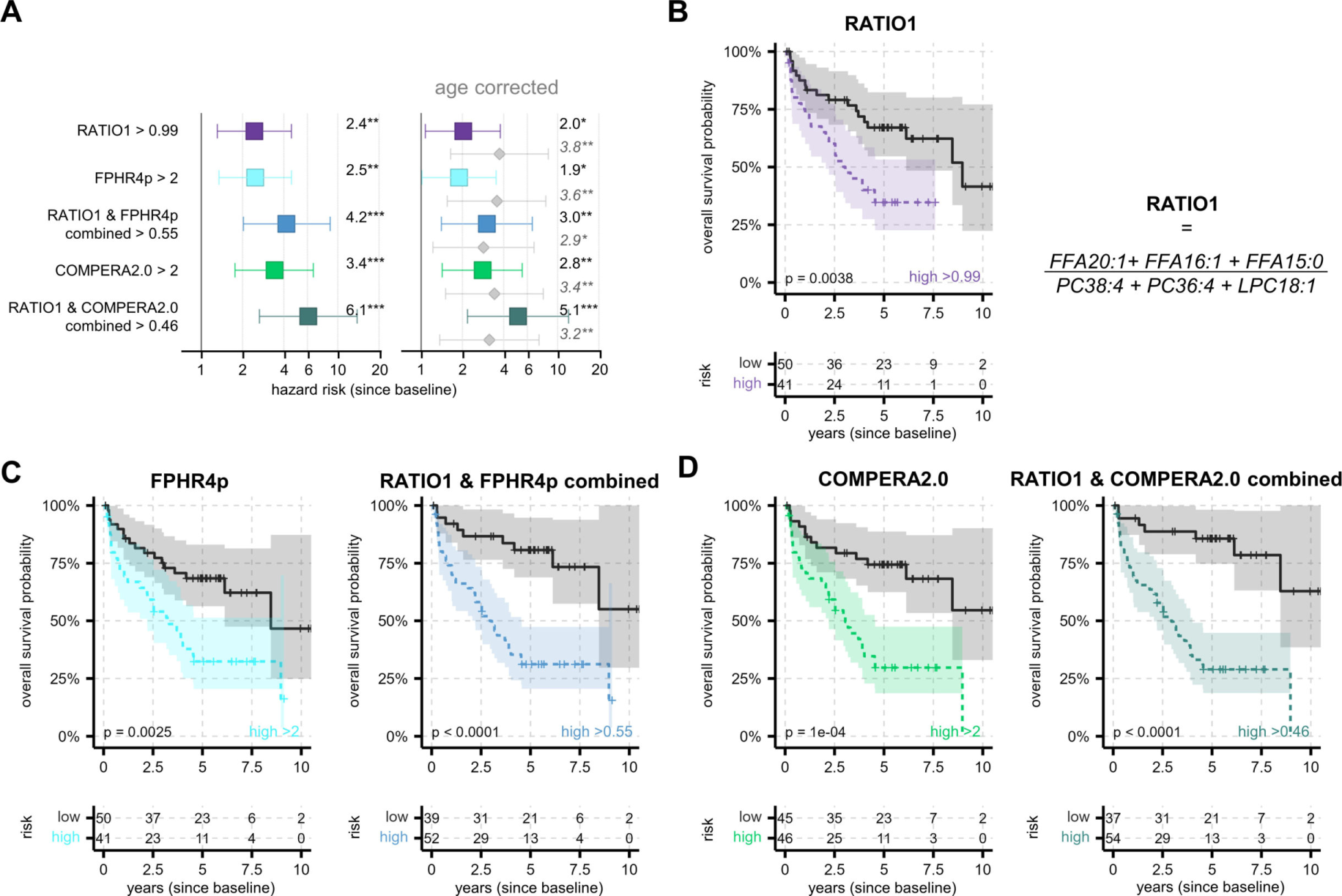
RATIO1 predicts survival and improves prediction of established clinical scores. **(A)** Cox HR analysis for survival from baseline without age as confounder (left side) and with age (right side, age HR shown in grey). Whiskers marks the 95% confidence intervals and statistical significance is coded as: * p < 0.05; ** p < 0.01; *** p < 0.001. Combining RATIO1 with FPHR4P or COMPERA2.0 increased HR compared with either alone. **(B, C, D)** Kaplan–Meier curves of survival from baseline by **(B)** RATIO1, **(C)** FPHR4p, and **(D)** COMPERA2.0 alone and combined with RATIO1. All cut-offs defining high or low were optimized with maxstat. RATIO1 was based on *log_10_*-transformed data without drift correction. RATIO1 was combined scaled 0 to 1 with equal weighing with either each score. FPHR4p was inverted so that higher scores represent higher risk as in COMPERA2.0.

As expected, established prognostic risk factors from the literature, higher WHO FC, lower 6MWD, and higher NT-proBNP were associated with poorer survival (Fig. S7). Next, we examined the major potentially confounding factors age, sex and BMI. Age > 57 yr constituted a considerable risk factor, while BMI>26.8 kg/m^2^ and male sex were not significant (Fig. S6). Results were similar in the joint model for all three factors, suggesting that only age was a relevant confounder. Therefore, we included only age as covariate to our HR analysis (Fig. 4A).

Overall, the HR results with age as covariate were similar to those without (Fig. 4A) and although age correction caused a decrease of all HR values, their respective prognostic impact remained significant.

### Changes in lipid metabolism in pulmonary arteries of idiopathic PAH (IPAH) patients

We investigated small PA from IPAH patients, the prototype of PAH and healthy donor lungs from explanted lungs. Oil red O staining and co-staining with markers of endothelial and smooth muscle cells showed accumulation of lipids in several IPAH PA (Fig. 5A). The observed lipid deposition could be the result of increased fatty acid uptake, metabolic dysregulation, or increased lipid synthesis. Therefore, we performed laser-capture microdissection of small PA (< 500 µm) from IPAH patients and healthy donors and examined gene expression of transporters and enzymes related to lipid metabolism. Gene expression showed significant upregulation of several key genes involved in lipid uptake and metabolism in IPAH (Fig. 5B). Most striking was the significant upregulation of SLC27A5, GAPT1, AGAPT1, Lipin2 and DGAT1. DGAT1 plays a critical role in lipid droplet formation. Up-regulation of the FFA transport protein SLC27A5 indicates increased uptake of FFAs from the circulation. GPAT, AGPAT, and lipin family enzymes promote triglyceride biosynthesis, incorporation of exogenous fatty acids into triacylglycerides (TAG) and phospholipids, as well as β-oxidation. The upregulation of these genes in the small PAs of IPAH patients might be caused by the increased circulating FFA levels or might be a manifestation of an underlying disease mechanism.

**Fig. 5.**
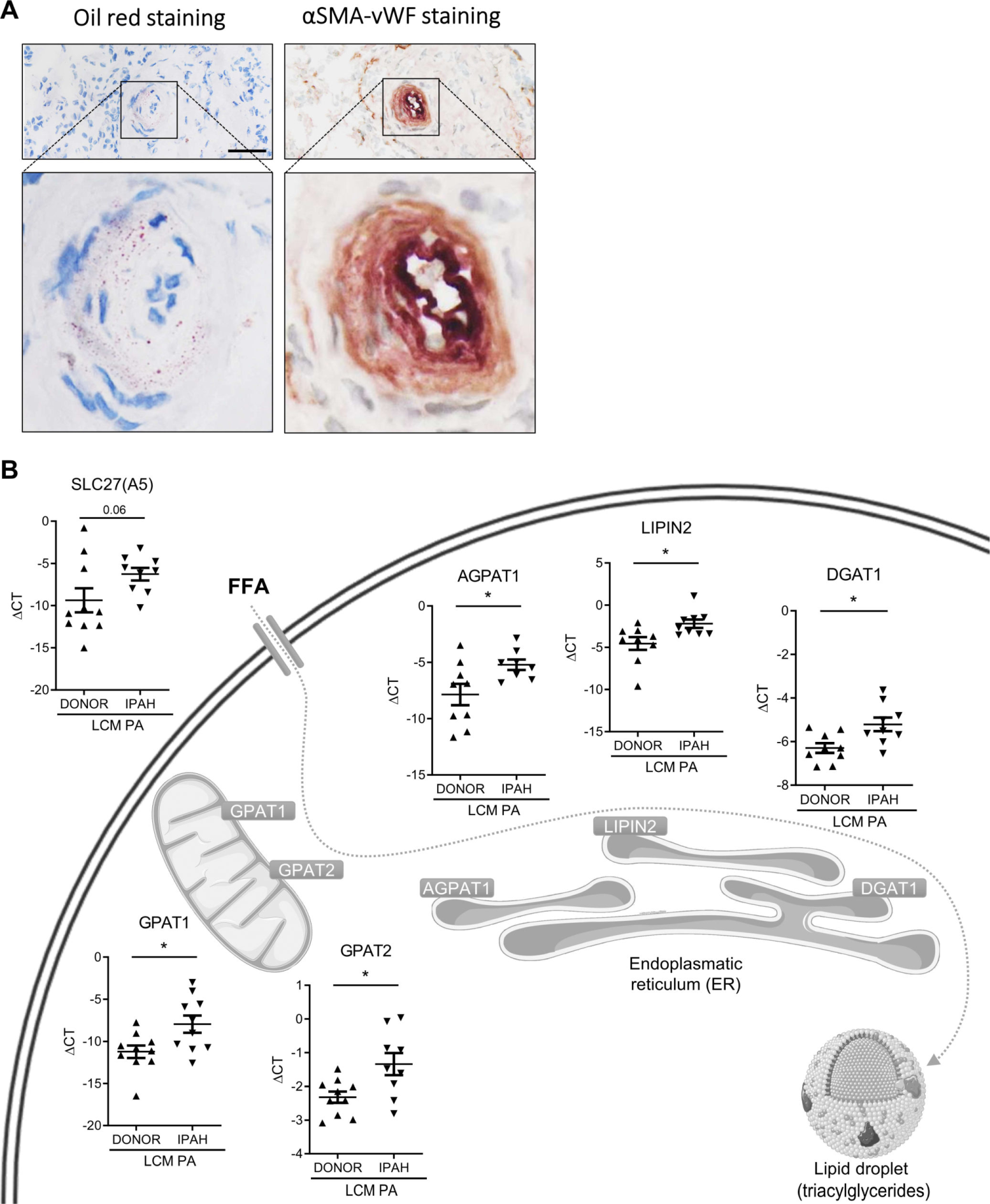
Presence of lipids and expression of lipid-metabolism genes in IPAH pulmonary arteries. **(A)** Visualization of endothelium (von-Willebrand factor, VWF) and smooth muscle cell layer (smooth muscle actin, SMA) together with oil red staining in human IPAH lung serial sections (scale bar: 50 µm). **(B)** Gene expression of lipid homeostasis-related enzymes in laser-capture microdissected human PA (n = 8-10 patients, respectively). Vertical lines represent means with standard error of mean (SEM). Asterisks mark Mann Whitney test p < 0.05. Cell organelles are adapted from Servier Medical Art (CC BY 2.0 DEED).

Next, we mimicked elevated circulating FFA levels by treating primary human pulmonary artery smooth muscle cells (hPASMC) and human pulmonary artery endothelial cells (hPAEC) from healthy donors with a FFA cocktail. Bodypi fluorescence staining showed accumulation of fat in hPASMCs and hPAECs (Fig. 6A). To better understand the functional effects of this, we performed in-vitro studies with primary hPAEC and hPASMCs. In hPASMCs, the treatment with the FFA mixture significantly promoted cell proliferation (Fig. 6B), and in hPAECs it significantly decreased acetylcholine (ACh)-induced NO secretion, suggesting endothelial dysfunction (Fig. 6C).

**Fig. 6.**
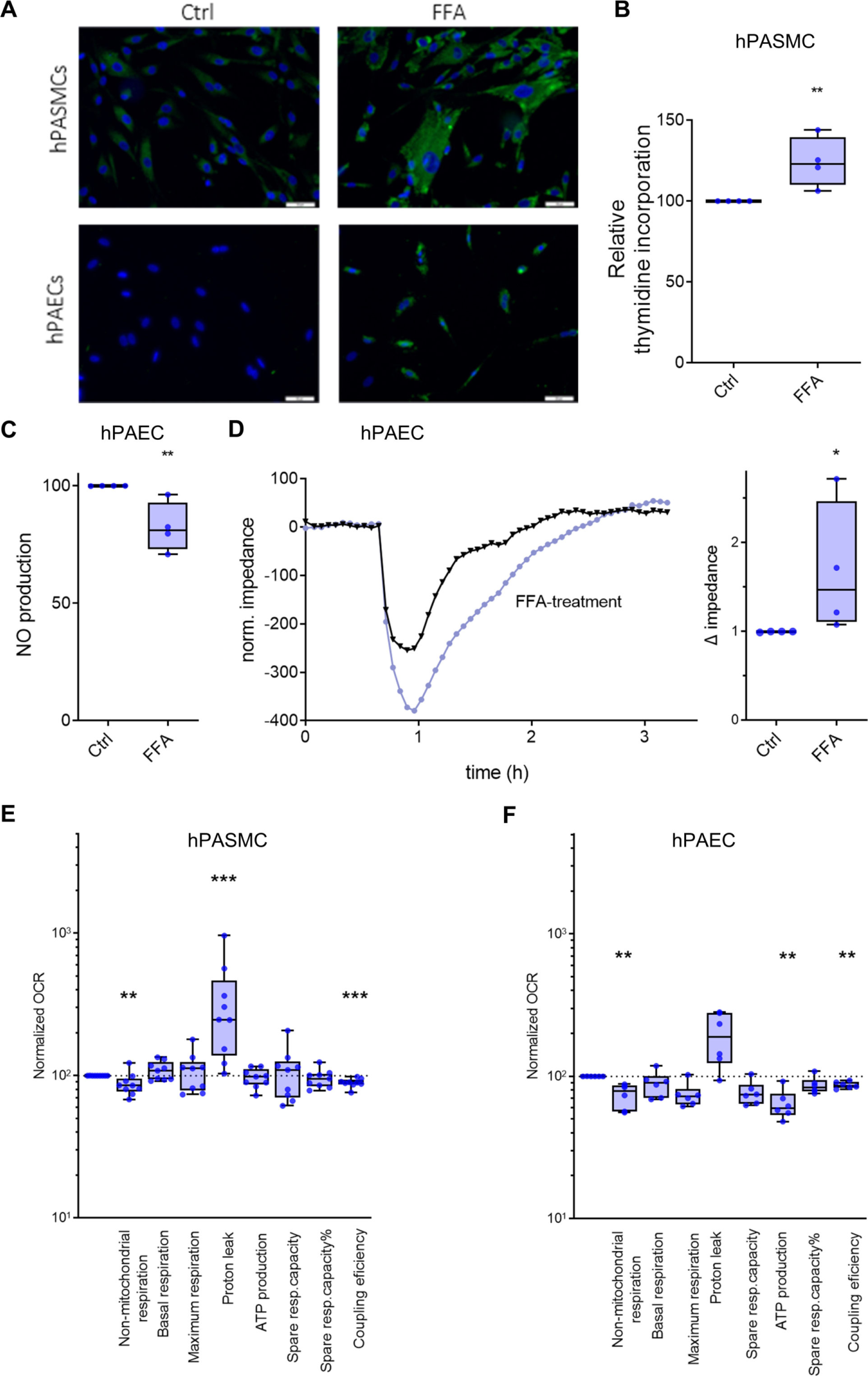
Effects of FFA treatment on hPASMC and hPAEC. **(A)** Representative Bodipy fluorescence staining of hPASMC and hPAEC in the absence (Ctrl) or presence of extrinsic FFA (scale bar: 50 µm). **(B)** Platelet-derived growth factor (PDGF)-BB induced proliferation of primary hPASMC measured with thymidine incorporation, in the absence (Ctrl) or presence of extrinsic FFA (n = 4). Changes are expressed as percentages compared with untreated controls (Ctrl). **(C)** ACh-induced NO production in primary hPAEC (n = 4). Changes are expressed as percentage compared with untreated controls (Ctrl). **(D)** TEER, as determined by electrical cell-substrate impedance sensor (ECIS), showed a significant decrease in hPAEC treated with FFA, suggesting endothelial leakage. Representative original curve (left panel) and changes expressed as percent change compared with controls (Ctrl) (n = 4). **(E, F)** Summarized data from hPASMC and hPAEC using the Seahorse XFe24 Extracellular Flux Analyzer. All measurements were performed on n = 25,000–50,000 cells/well and five wells per cell type. Each experimental group consisted of cell lines from two to three patients. All data were normalized to total protein per well before analysis (n = 9). Mann Whitney test * < 0.05, **< 0.01, ***< 0.001. The boxes extend from the 25^th^ to 75^th^ percentile, the middle line denotes the median and the whiskers mark the minimum and maximum.

We investigated the effect of the FFA mixture on endothelial barrier function by determining the magnitude of thrombin-induced endothelial barrier dysfunction. Fig. 6D shows the typical response of control endothelium to thrombin. When endothelial monolayers were pre-treated with FFA, they exhibited a significantly pronounced decrease in transendothelial electrical resistance (TEER) and delayed recovery of barrier function compared to control media (Fig. 6D). This suggests that FFA treatment causes profound endothelial dysfunction. Finally, we examined FFA-induced metabolic responses in hPASMCs and hPAECs by means of the Seahorse method and found significantly decreased coupling efficiency in both cell types in response to FFA treatment (Fig. 6E, F). Moreover, FFA exposure decreased non-mitochondrial respiration and ATP production in hPAEC and increased proton leak in hPASMC. This suggests that FFA treatment changes the phenotype of hPASMCs and hPAECs from healthy donors into an IPAH phenotype and that high levels of circulating FFAs may cause pulmonary vascular dysfunction, representing either a primary cause or a novel vicious circle in PH.

## Discussion

The uptake and metabolism of long-chain fatty acids is critical for many physiological and cellular processes, and cellular accumulation may cause numerous pathological and functional changes. Previous investigations in PAH have shown severe metabolic changes of the right ventricle and elevated *in vivo* myocardial triglyceride content^10,22,23^. Our investigations add important information by showing that small PA of IPAH patients are affected by lipid accumulation. Interestingly, we found increased gene expression of enzymes causing fatty acid uptake and triglyceride biosynthesis in smooth muscle cells of IPAH patients (Fig. 5C). This could be caused by the high FFA levels in the circulation, however, it could also represent a change in the cell physiology that strongly contributes to the development of PH.

For the first time, we have explored the effects of FFA exposure in primary human hPAECs and hPASMCs. FFA exposure decreased NO secretion and impaired barrier function in hPAECs, and caused increased proliferation in hPASMC. In addition, FFA exposure induced changes in non-mitochondrial respiration and coupling efficiency in both cell types. This suggests that impaired lipid handling in IPAH PAs might trigger the remodelling in PAH. This is in line with numerous studies indicating that the expression of GAPT1 / AGPAT1 / lipin-1 has important metabolic consequences^24–27^. Our data, taken in context with data from the literature, may suggest that in PH, the failing right ventricle is no longer able to cope with FFA metabolism, leading to an increase in circulating FFA levels, which negatively affects the pulmonary vessels. However, it is also possible that there are primary changes in the lipid metabolism of the small PAs, leading to vascular dysfunction, subsequently increasing right ventricular afterload, initiating a vicious circle that finally causes PH and right heart failure.

Spanning over two decades, extensive basic, translational, and clinical analyses have supported a causative link between metabolic reprogramming and PAH^28,29^. Similar to the Warburg effect in cancer, a shift from mitochondrial oxidation to glycolysis appears to occur in the right ventricle of PAH patients^22,23^. In this study, we show that the small PAs are affected by significant metabolic changes. Taking cues from cancer, recent data demonstrate significant alterations in metabolic programs other than glycolysis and glucose oxidation, including the pentose phosphate pathway (PPP), glutaminolysis, lipolysis, fatty acid synthesis and oxidation and changes in the plasma proteome ^10–16,30–32^. However, it remains unclear whether these changes originate in the overloaded right ventricle, in the primarily affected pulmonary vessels, or elsewhere.

Although there has been tremendous progress in the understanding of PAH in recent decades, there is still an unmet need for diagnostics and therapy. According to a recent literature review, the five-year survival rate for newly diagnosed patients has not significantly improved despite a multitude of new PAH medications^8^. This may be due to the fact that the metabolic mechanisms of the disease have not been addressed in detail. To our knowledge, this study is the first to apply unsupervised, broad metabolome analysis of PH patients of group 1 to 4. The generated specific FFA/lipid-ratios identified patients with PH, independent of comorbidities. The same ratios provided prognostic information, complementing existing clinical prognostic scores. The diagnostic and prognostic results were validated in an independent international cohort and this was confirmed by a balanced split group approach. Of note, our *in vitro* mechanistic studies suggest that disturbed lipid metabolism may significantly contribute to the pathologic mechanisms in IPAH patients. Our simple FFA/lipid-ratios might be useful in the diagnosis and clinical management of PH patients and might even serve as surrogate endpoints in future clinical trials.

Classical machine learning using RF and XGboost showed that metabolic differences hold a high potential for diagnostic biomarkers. Both approaches were able to overcome typical technical MS-specific problems such as batch effects and intensity jumps between measurements, which usually require drift correction, suggesting that such labour-intensive procedures are not necessary if PH is to be detected. The same is true for our easily applicable FFA/lipid-ratios that performed comparably well with drift-corrected and non-corrected metabolomics data, suggesting that forming such a ratio corrects for batch effects and drifts in the metabolomics dataset as well as for inter-individual variability of patients’ lipid metabolism and lifestyle.

Our FFA/lipid-ratios performed very well in both PH diagnosis and survival prediction. Their diagnostic performance even outperformed RF and XGBoost models, especially in terms of sensitivity. In addition, the results were stable despite the use of different sample types such as serum, heparin and EDTA plasma from three different centers, a very important prerequisite for broad applicability in routine diagnostics. The performance was also stable when training and validation cohort were not split by center but 70% to 30% balanced by age, BMI, sex and class. The FFA/lipid-ratios are easy to measure and fully explainable compared to machine learning models, which is advantageous for future studies and regulatory approval processes for *in vitro* diagnostics (IVD).

Survival prediction is an important tool in the management of PAH patients. Several clinical scores have been established, with FPHR4P and COMPERA2.0 representing most recent developments derived from large databases^20,21^. The FPHR4P^20^ score estimates prognosis based on WHO FC, 6MWD, CI, and RAP, while COMPERA2.0^21^ is based on non-invasive parameters, only (WHO FC, 6MWD, NT-pro-BNP). RATIO1 showed an age-dependency that was comparable to both clinical scores. Most importantly, both clinical scores gained notably in predictive power when combined with RATIO1. This suggests that FFA/lipid-ratios represent an independent, non-invasive prognostic factor that combines favourably with established prognostic PH scores.

### Strengths and limitations

Strengths of this study include exploration of primary IPAH small PA vessels, with mechanistic insight in the effects of FFA on PASMC and PAEC, a broad metabolomics approach, sampling and processing conditions suitable for routine clinical practice, inclusion of a disease control group, comprehensive clinical assessment, use of machine learning, and development of diagnostic and predictive FFA/lipid-ratios. As a limitation, we had access to a small number of patients and controls. This may have been compensated by a profound clinical characterization of the patients, including RHC, coupled with long follow-up times for survival analysis and the 70% to 30% balanced split test, confirming the results. Another limitation is that FPHR4p and COMPERA2.0 have been derived from PAH patients while we used them for all available patients including PH associated with left heart and lung diseases. This may have introduced a bias into the prognostic performance of these scores, however, this bias relates to the scores and our new markers in the same way. It may be seen as a limitation, that blood samples were collected along with clinical routine blood draws, without standardized fasting or other control measures, however, this may also represent a strength of our study as it suggests robustness of the results. All metabolic measurements were based on high-resolution mass spectrometry, a very sensitive and exact method, yet slow, expensive and work-intensive. However, our FFA/lipid-ratios allow for a simplified approach that is easily available.

### Outlook

Future studies including larger numbers of patients with a balanced group distribution and more patients with early pulmonary hypertension and less impaired right ventricular function and longitudinal studies are warranted to investigate the value of metabolic markers for patient management.

### Conclusions

Based on our mechanistic insights into the metabolic changes in small PA, and our machine learning approaches, our FFA/lipid-ratios identified PH patients with a high accuracy and were significantly associated with prognosis. This may point to novel diagnostic tools and possibly also to new therapeutic targets. If implemented into the management strategy for PAH patients, this might inform therapy decisions to improve outcomes of PAH therapy.

## Supplemental Information

Supplemental Information can be found attached to this publication.

## Materials Availability

This study did not generate new unique reagents.

## Data and Code Availability

The author declare that all data supporting the findings in this study are available in the online supplementary data 1 and online repositories. Mass spectrometric data have been deposited in https://zenodo.org under doi: 10.5281/zenodo.7857706. Data is provided de-identified and is available immediately after publication with no end for those who wish to access the data for any purpose. The provided *Sample_Name* in the online supplementary data 1 links to the file names in the online repository and are unsuitable to identify single patients. The primary key is only known to part of the study team. Machine learning code is available immediately with no end for those who wish to access for any purpose on Github: https://github.com/HelgaLudwig/PHMetab.

## Author contributions

Conceptualisation, NB, BMN, TP, HO, AO; Data curation, NB, BMN, EZ, HL, UB, HO, AO; Formal analysis, NB, BMN, EZ, HL, UB, HO, AO; Funding acquisition, NB, TP, HO, AO; Investigation, NB, BMN, EZ, VF, CN, VB, HO, AO; Methodology, NB, BMN, EZ, CN, AO, HO; Project administration, NB, BMN, UB, VF, KH, SU, TJL, HO, AO; Resources, NB, BN, EZ, CM, HL, UB, KH; SU, TJL, TP, HO, AO; Software, NB, HL; Supervision, NB, EZ, UB, HO, AO; Validation, NB, BN, EZ, HL, VF, CN, VB, SU, TJL, HO, AO; Visualisation, NB, BMN, HL, CN, VB, AO; Writing – original draft, NB, BAM, HO, AO; Writing – Review & Editing, all authors;

## Data Availability

The author declare that all data supporting the findings in this study are available in the online Supplementary Sata 1 and online repositories. Mass spectrometric data have been deposited in https://zenodo.org under doi: 10.5281/zenodo.7857706. Data is provided de-identified and is available immediately for any purpose. The provided Sample_Name in the online Supplementary Data 1 links to the file names in the online repository and are unsuitable to identify single patients. The primary key is only known to part of the study team. Machine learning code is available immediately for any purpose on Github: https://github.com/HelgaLudwig/PHMetab.

https://zenodo.org/records/7857706

https://github.com/HelgaLudwig/PHMetab

## Abbreviations

6MWD: six minute walking distance
ACh: acetylcholine
ADMA: asymmetric dimethylarginine
AUC: area under the curve
BH: Benjamini–Hochberg
BMI: body mass index
BNP or NT-proBNP: natriuretic peptide levels
CO: cardiac output
COPD: chronic obstructive pulmonary disease
CI: cardiac index
CTEPH: chronic thromboembolic pulmonary hypertension
DC: diseased control (non-PH)
DLCOcVA: diffusing capacity for carbon monoxide per alveolar volume, hemoglobin corrected
ECAR: extracellular acidification rate
ECIS: electrical cell-substrate impedance sensor
EDTA: ethylenediaminetetraacetic acid
FEV1: forced expiratory volume/ 1 s
FVC: forced vital capacity
FFA: free fatty acids
H&E: hematoxylin-eosin
HC: healthy control
HILIC: hydrophilic interaction liquid chromatography
hPAEC: human pulmonary artery endothelial cells
hPASMC: human pulmonary artery smooth muscle cells
HR: hazard ratio
HRMS: high resolution mass spectrometry
ILD: interstitial lung disease
IPAH: idiopathic pulmonary arterial hypertension
iPCA: independent principal component analysis
IVD: *in vitro* diagnostics
LPC: lysophosphatidylcholine
LPE: lysophosphatidylethanolamine
LV: left ventricle
MAD: median absolute deviation
mPAP: mean pulmonary arterial pressure
MS: mass spectrometry
OCR: oxygen consumption rate
OPLS-DA: orthogonal projections to latent structures discriminant analysis
PA: pulmonary arteries
PAH: pulmonary arterial hypertension
PAP: pulmonary arterial pressure
PAWP: pulmonary arterial wedge pressure
PBS: phosphate-buffered saline
PC: phosphatidylcholine
PDGF: platelet-derived growth factor
PH: pulmonary hypertension
PPP: pentose phosphate pathway
PVR: pulmonary vascular resistance
QC: pooled from samples for quality control
RAP: right atrial pressure
RDW: red cell distribution width
RF: random forest
RHC: right heart catheterization
ROC: receiver operator curve
RT: room temperature (20 – 25 °C)
SEM: standard error of mean
SM: sphingomyelin
SMA: smooth muscle actin
SvO2: mixed venous oxygen saturation
TAG: triacylglyceride
TEER: transendothelial electrical resistance
TLC: total lung capacity
VWF: von-Willebrand Factor
WHO FC: World Health Organization functional class
WU: Wood unit
XGBoost: eXtreme Gradient Boosting

## Acknowledgements

We are very grateful for the excellent technical assistance from Elisabeth Blanz, Sabine Halsegger, Daniela Kleinschek, Jessica Schweiger, Yasemin Gassner, Gert Trausinger and Edgar Gander. We express our heartfelt gratitude to Gabor Kovacs, Saskia Trescher, Pablo López-García, Sophie Narath, Michael Pienn and Peter Wolf for their valuable discussions and helpful advices.

## Online Methods

### Cohort Data Sources

Results from the Graz Pulmonary Hypertension Registry (GRAPH) have been reported previously^1,2^. Briefly, the program uses a software application linked to the electronic health record for documentation of all patients of the Division of Pulmonology at the Department of Internal Medicine of the Medical University of Graz who gave written informed consent. Demographic, clinical, echocardiographic, procedural, and hemodynamic data and blood samples are collected and tracked for longitudinal outcomes. All hemodynamics were measured in a standardized fashion by the same experienced team^1^. Regularly scheduled quality checks of the registry are performed to ensure completeness and accuracy. Written informed consent was obtained from all patients and the study was conducted in line with the Helsinki declaration. The study was approved by the institutional ethics board (identifier: 23-408ex10/11), and the study has been registered at ClinicalTrials.gov (NCT01607502).

The BioPersMed (Biomarkers of Personalised Medicine) project is designed as a single-centre, prospective, observational cardiovascular risk study. Between 2010 and 2016, 1022 community dwelling and asymptomatic individuals were regionally recruited and assessed biannually including a standardized biospecimen acquisition^3^. Written informed consent was obtained from all patients and the study was conducted in line with the Helsinki declaration.

### Cohort Study Population

We retrospectively enrolled two consecutive cohorts of patients identified through our single-center GRAPH registry and labelled them as GRAPH-Metabolomics (GRAPH-M).

The inclusion criteria for the first cohort were diagnosis of idiopathic pulmonary arterial hypertension (IPAH) and absence of severe co-morbidities. Healthy sex- and age-matched subjects served as a healthy controls (HC).

Inclusion criteria for the second cohort consisted of the following PH groups 1 – 4: 1) PAH, or 2) PH associated with heart disease, 3) PH associated with lung diseases (COPD, ILD), or 4) CTEPH. The cohort of disease controls (DC) consisted of patients with airway or parenchymal lung disease or patients with metabolic syndrome (hypertension, hypercholesterinemia and type 2 diabetes mellitus) but no signs of elevated PAP. Healthy sex- and age-matched subjects served as a control group (HC). The patients with the metabolic syndrome were selected from the BioPersMed cohort Graz.

All cohorts with the exception of patients with metabolic syndrome or HC underwent RHC. In patients who underwent multiple RHCs, the first RHC was considered the index procedure and was the only one included in the analysis. Patients were included in the analyses if data from a complete RHC were available, including a resting value for mPAP, PAWP, CO, heart rate, systolic and diastolic PA pressure, PVR and mixed venous oxygen saturation (SvO_2_). We used the standard equation to calculate the pulmonary vascular resistance with PVR = (mPAP - PAWP)/CO expressed in WU.

### Validation Cohort

This cohort comprised an international multicentric patient cohort. The inclusion criteria for the validation cohort were the confirmed diagnosis of PAH and informed written consent at the home institution. All participating centers were experienced centers of excellence for PH and all patients underwent RHC in a standardized manner. Demographic, clinical, echocardiographic, procedural, and hemodynamic data and blood samples were available as anonymized data. The sample collection was approved by the local Ethics Committee in each local center (Regensburg University, ethics committee No. 08/090 and cantonal ethical review board Zürich KEK 2010-0129; 2014-0214; 2017-0476).

### Cohort Outcomes

The primary outcome was the confirmation of PH, defined as mPAP ≥ 25 mmHg according to the ERS/ESC guidelines from 2015^4^. The secondary endpoints included time to all-cause mortality with data provided by Statistic Austria (single-centre registry Graz) and by the respective centers who contributed to the validation cohort. A complete list of covariates analyzed in this study is provided in the Supplementary Material (Fig. S3).

### Human lung tissue samples

Human lung tissue samples were obtained from patients with IPAH who underwent lung transplantation at the Department of Surgery, Division of Thoracic Surgery, Medical University of Vienna, Vienna, Austria. The protocol and tissue usage were approved by the institutional ethics committee (976/2010) and written patient consent was obtained before lung transplantation. The patient characteristics included: age at the time of the transplantation, weight, height, sex, mPAP measured by RHC, pulmonary function tests, as well as the medical therapy. The chest computed tomography scans and RHC data were reviewed by experienced pathologists and pulmonologists to verify the diagnosis. Healthy donor lung tissue was obtained from the same source. Donor/IPAH patient characteristics are given in Supplementary Material (Table S2)**Fehler! Verweisquelle konnte nicht gefunden werden.**.

### Lung histological Oil Red O staining

The lipid accumulation of lung tissues was visualized by Oil Red O staining (Merck KGaA, Darmstadt, Germany). Lung tissue was embedded in tissue freezing medium, were snap frozen at −80 °C and sliced using a Leica CM 1900 cryostat (Leica Biosystems GmbH, Wetzlar, Germany) at a thickness of 5 μm per section. The slices were stained with Oil Red O working solution for 10 min, differentiated in isopropanol for 5 min, and then washed with water at room temperature (20 – 25 °C) (RT). The experiments were finished according to the manufacturer’s instructions. The morphological features of the tissues were assessed by hematoxylin-eosin (H&E) staining. The lipid in the tissue observed by microscope.

### Laser capture microdissection of PA and RNA extraction

Laser capture microdissection (LCM) of 10 donor lungs and 10 lungs from IPAH patients, as well as mRNA isolation and cDNA synthesis were performed as previously described^5^. The intima and media layers of PAs of 100 – 500 μm diameter were selected, marked and isolated with the Arcturus^®^ LCM System. Captured vessels were immediately transferred into RNA lysis buffer and were snap frozen. RNeasy Micro Kit was utilized to isolate RNA (RNeasy Micro Kit, Qiagen, Hilden, Germany)^6^.

### qRT-PCR - laser capture microdissected human PA

The expression of enzymes and transporters was analyzed with real-time quantitative (qRT)- PCR using the QuantiFast SYBR PCR reagent (Qiagen, Hilden, Germany) according to Papp et al. 2019^7^. Primer pairs (Eurofins, Graz, Austria), summarized in Supplementary Material (Table S3), were designed to span at least one exon-exon boundary to avoid the amplification of genomic DNA. The specificity of all primers, as well as the length of the amplicon, were confirmed by melting curve analysis and by running the products on 2% agarose gels, respectively.

### Cell Isolation and culture

#### hPAECs

hPAECs were either purchased from Lonza or isolated from donor lungs. For the isolation of donor hPAECs, PA (< 2 mm in diameter) were isolated and the endothelium incubated with an enzymatic mixture of collagenase, DNAse and dispaze in HBSS at RT^8^. Cell suspension was collected, resuspended in VascuLife Complete SMC Medium and cultured in gelatin-coated T25 flasks at 37°C and 5% CO_2_. After reaching 70 – 80% confluency, cells were trypsinized, enriched by 3 consecutive steps of CD31-selective magnetic-activated cell sorting technology and verified via morphological and marker confirmation (smooth muscle actin SMA, fibronectin, vimentin, von-Willebrand Factor VWF, smooth muscle myosin heavy chain and CD31). Surplus hPAECs were frozen (endothelial cell complete medium containing 12% FCS and 10% DMSO) and stored in liquid nitrogen until further use. Passages 2–6 were used for the experiments. Detailed patient characteristics of isolated hPAECs can be found in Supplementary Material (Table S4).

#### hPASMCs

The isolation and culture of human hPASMCs was performed as previously reported^9^. After the removal of the endothelial cell layer, the media was peeled away from the underlying adventitial layer and cut into approximately 1–2 mm^2^ sections, centrifuged and resuspended in VascuLife Complete SMC Medium supplemented with 20% FCS and 0.2% antibiotics, then transferred to T75 flasks and cultured at 37°C and 5% CO_2_. After confluency of hPASMC was formed, the cells were trypsinized and either cultured in VascuLife Complete SMC medium supplemented with 10% FCS and 0.2% antibiotics, or frozen (VascuLife Complete SMC Medium containing 15% FCS and 10% DMSO) and stored in liquid nitrogen until further use. Passages 4–8 were used for the experiments. SMCs were verified via morphological and marker confirmation (smooth muscle actin SMA, fibronectin, vimentin, von-Willebrand Factor VWF, smooth muscle myosin heavy chain and CD31). Detailed patient characteristics of isolated hPASMCs can be found in Supplementary Material (Table S4).

### Lipid visualized by bodipy staining

BODIPY (D3922, Invitrogen, Carlsbad, Calif, USA) (excitation wavelength 493 nm, emission maximum 503 nm), was diluted in phosphate-buffered saline (PBS, 137 mM NaCl, 2.7 mM KCl, 12 mM HPO ^2−^/H PO ^−^, pH 7.4) or DMSO at a concentration of 1 mg/mL and applied to the hPASMCs and hPAECs for 20 mins at RT. Fixed cells (4% paraformaldehyde at 37°C for 5 mins) were used. Following fixation, samples were washed 3 times in PBS for 10 min. Sections were counterstained with 4′,6-diamidino-2-phenylindole dihydrochloride (DAPI; Sigma-Aldrich) to visualize nuclei and covered with glass cover slips. Images were taken using a laser scanning confocal microscope (Zeiss LMS 510 META; Zeiss, Jena, Germany) with Plan-Neofluar (×40 /1.3 Oil DIC) objective.

### Measuring cell metabolic state

Oxygen consumption rate (OCR) and extracellular acidification rate (ECAR) were determined by the Seahorse XFp analyzer (Agilent, USA)^10^. hPASMCs or hPAECs were plated onto cell culture microplates on the day before the experiments and treated with 0.25mM FFA (a mix of oleate, FFA 16:0 and FFA C18 (2:1:1)) in VascuLife® complete Medium and incubated for 24 h. Cells were then incubated in XF assay medium (Agilent), supplemented with 25 mmol/L glucose and 1 mmol/L pyruvate (hPASMCs) or 10 mmol/L glucose, 1 mmol/L pyruvate, and 2 mmol/L L-glutamine (hPAECs) for 1 h at RT before the measurement. After the recording of the basal rates of OCR and ECAR, final concentrations of 1 μmol/L oligomycin, 1 μmol/L carbonyl cyanide-4 (trifluoromethoxy) phenylhydrazone, and 0.5 μmol/L rotenone and antimycin A for human hPASMCs; 1 μmol/L oligomycin, 1 μmol/L carbonyl cyanide-4 (trifluoromethoxy) phenylhydrazone, and 1 μmol/L rotenone and antimycin A for human hPAECs were added (XF Cell Mito Stress Test Kit, Agilent) through the instrument’s injection ports to obtain proton leak, maximal respiratory capacity, and nonmitochondrial respiration, respectively. Glycolytic capacity was measured using an XF Glycolysis Stress Test Kit (Seahorse Bioscience). ECAR was determined after serial injection with 10 mmol/L D-glucose, 1 μmol/L oligomycin, and 100 mmol/L 2-deoxyglucose. All the assays were performed in triplicate and normalized to protein content.

### Endothelial Barrier Function

TEER served as an indicator of barrier function of endothelial cell monolayers. TEER was determined using an electrical cell-substrate impedance sensor (ECIS) (Applied Biophysics, Troy, NY, USA). Briefly, the endothelial cells are seeded in complete medium into (8W10E -PET arrays Applied Biophysics, NY, USA) each well and allowed them to grow until they reached confluence. The FFA (0.25 mM, a combination of FFA 18:1, FFA 16:0 and FFA 18:0 (2:1:1)) was applied for 24 hours before the barrier disruption was initiated by addition of recombinant human thrombin.

### Proliferation

To investigate the proliferative effect of FFA treatment on hPASMCs, the following protocol was applied^11^: 10 000 hPASMCs were seeded in 96-well plates; the following day the cells were starved (VascuLife® Basal Medium, 0% FCS, 0.2% antibiotic/antimycotic) or kept under control conditions (VascuLife® Basal Medium with 5% FCS; LifeLine Technology, Walkersville) for 12 h. Afterwards, platelet-derived growth factor (PDGF)-BB was added and the proliferation of hPASMCs was determined by [^3^H] thymidine (BIOTREND Chemikalien GmbH) incorporation, after 24 h of incubation, as an index of DNA synthesis and measured as radioactivity by scintillation counting (Wallac 1450 MicroBeta TriLux Liquid Scintillation Counter and Luminometer). To investigate the effect of FFA (0.25 mM, a combination of oleate, FFA 16:0 and FFA 18:0 (2:1:1)) on hPASMCs, the same number of cells was seeded and after 12 h of starvation, FFA and vehicle were added and the proliferation of hPASMCs was determined as aforementioned. All experiments were performed in quadruplicate.

### DAF-DM-mediated nitric oxide measurement

Measurements were performed as previously described^8^. hPAECs were seeded in gelatin-coated dark 96-well plates, starved for 2 h with Ringer’s solution and loaded with 10 µm 4-Amino-5-Methylamino-2′,7′-Difluorofluorescein Diacetate (DAF-FM) for 30 min at 37°C. The cells were stimulated with 5 µM acetylcholine (ACh) for the induction of nitric oxide measurement on CLARIOstar Plus (BMG Labtech, Ortenberg, Germany) at Ex/Em = 495/515 nm. All the assays were performed in quadruplicate and normalized to protein content.

### Plasma liquid chromatography–mass spectrometry metabolomics

Metabolites were analysed by targeted hydrophilic interaction liquid chromatography–high resolution mass spectrometry (HILIC-HRMS) metabolomics according to Bajad et al.^12^ and samples were processed according to Yuan et al.^13^ as described previously^14,15^.

Samples from Graz were aliquoted and stored at the Biobank Graz. On the day of the processing they were thawed in water ice bath on a slow rotary shaker (300 rpm) in < 10 min and vortexed shortly. Aliquots of 100 µl were precipitated in LoBind Eppendorf tubes with 400 µl cold methanol (-80°C for at least 4 h, kept on dry ice) and vortexed shortly. After the overnight precipitation at –80 °C the samples were centrifuged for 10 min at 14.000 g at 4°C and supernatants transferred to fresh LoBind Eppendorf tubes. Supernatants were dried under nitrogen flow and stored at –80°C until all batches of the cohort were finished. Extracts were reconstituted in 100 µl 30% methanol/H_2_O, vortexed for 45 s and centrifuged for 5 min at 14.000 g at 4°C. The supernatant was transferred into autosampler vials, and equal aliquots from all samples were pooled for quality control (QC). All ready-to-measure extracts were refrozen at –80°C prior to measurement. Every 24 h samples were freshly thawed at RT, vortexed, spun down and added to the autosampler at 4°C.

Measurements were made in independent runs per cohort with samples in randomized order, interspaced by according blanks, pooled QC samples and UltimateMix (UM, described previously^16^). Pooled QC samples were generated independently for cohort 1 and 2, while QC was mixed for cohort 3 and 4. Cohorts 1 and 2 were extracted and measured in 1 batch, while cohorts 3 and 4 were randomly divided into 6 batches for sample extraction and measured with daily thawed extracts to reduce metabolite degradation.

Extracts were measured with a Dionex Ultimate 3000 high-performance liquid chromatography (HPLC) setup (Thermo Fisher Scientific, USA) equipped with a NH_2_-Luna HILIC analytical column and crudcatcher with an injection volume of 10 µl and a 37 min gradient from aqueous acetonitrile solution [(5% acetonitrile v/v), 20 mM ammonium acetate, 20 mM ammonium hydroxide, pH 9.45] as eluent A (LMA) to acetonitrile as eluent B (LMB). Mass spectrometric detection was carried out with a Q-Exactive^TM^ system (Thermo Fisher Scientific). Electrospray ionization (ESI) was used for negative and positive ionization and masses between 70 and 1050 m/z were detected.

Raw data were converted to mzXML using msConvert (ProteoWizard Toolkit v3.0.5), and target metabolites were extracted using the in-house developed tool PeakScout. Spectrum slices were presented around the exact target mass (± 50 ppm) and retention time (± 3 min) in accordance with the standards described by Sumner et al.^17^. For each target metabolite all peak area integrations were manually confirmed in each sample. Molecular masses of target metabolites were taken from literature and available online databases (HMDB, KEGG, Metlin)^18–20^. In addition, pure substances of all hydrophilic metabolites and selected lipophilic metabolites were run on the same system to obtain accurate reference retention times and fragmentation spectra.

The analytical quality of all targeted metabolites was strictly graded to be suitable for multivariate analysis and univariate analysis using the following parameters: deviation from target mass < 5 ppm, mass difference range < 10 ppm, retention time standard deviation < 0.75 min, percentage of missing values < 30%, relative standard deviation in QC after drift correction <30%, and blank load in QC < 30%. Of 164 included metabolites, 11 metabolites were considered unsuitable for ROC analysis due to lower signal intensity and lower consistency in repeated sample measurements (controls in cohorts 3 and 4).

Analytical quality of samples, blanks, QC, and UM was graded by sample median, peak shapes, retention time shifts, percentage of missing values < 30%, and position in the PCA scores plot. From cohort 2, the first two UM and from cohort 4 the last four QC did not meet the quality criteria and were therefore excluded.

### Statistical analysis

Data visualisation and statistical analysis were performed with R v4.0.2 (R Core Team, 2020) (using the packages readxl, openxlsx, stringr, dplyr, tidyr, doParallel, statTarget, car, colorspace, RColorBrewer, ggplot2, ggforce, ggpmisc, ggpubr, scales, grid, ellipse, correlation, dendsort, pheatmap, nlme, emmeans, missMDA, FactoMineR, mixOmics, MetaboAnalystR 3.0.3, survival, survminer, pROC, caret, patchwork) and TIBCO Spotfire v12.5.0 (TIBCO, Palo Alto, CA). Graphpad Prism v9 has been used to assess differences in the *in vitro* experiments.

Typically, MS results are relative and only comparable within the same run. However, recent advances in drift correction allow to merge data for joint analysis. Peak areas without drift correction were *log_10_-*transformed prior to all further analysis. The drift correction was based on the RF driven algorithm that used QC measurements to model batch effects and drift for each metabolite with *statTarget::shiftCor(., Frule = 0.7, ntree = 500, impute = “KNN”, coCV = 100, QCspan = 0, degree = 2)*^21^. The imputed, drift-corrected data were multiplied by 10^3^ to make the numbers more readable after *log_10_-*transformation. In the drift corrected, *log_10_*-transformed data all imputed values were removed and data was trimmed by median absolute deviation (MAD) score^22^, assuming a normal distribution (multiplication with 1.4826). Strong single outliers were removed with a very conservative threshold of having an absolute MAD score > 4 (165 single values in 65 metabolites). Data for all metabolites and samples is provided in Supplementary Data 1 with and without drift correction.

In order to ensure validity of drift correction and subsequent results, each measurement run was first analysed independently with unsupervised multivariate (iPCA), supervised multivariate (OPLS-DA) and univariate on *log_10_*-transformed data without drift correction (see Fig. S1). Next, drift corrected, *log_10_*-transformed data from all runs was jointly analysed with the same methods. The drift correction successfully removed the significant difference between measurement runs and notably reduced the technical variability in all metabolites (Fig. S2). Additionally, no difference was observed based on center or sample material type (Fig. S2). Replicate measurements of samples in different runs were average on drift corrected data to yield one metabolome per patient for subsequent analysis one metabolome per patient.

All reported p-values were adjusted for multiple testing according to Benjamini–Hochberg (BH) denoted as p_BH_ (*stats::p.adjust()*)^23^. Distribution and scedasticity were investigated with Kolmogorov–Smirnov test (s*tats::ks.test()*) and Brown–Forsythe Levene-type test (*car::leveneTest()*)^24^, respectively. After *log_10_-*transformation data was mostly normally distributed with 91% of all metabolites without drift correction and 99% of all metabolites with drift correction testing not significant (p_BH_ > 0.05). Analog, data was mostly homoscedastic with 79% without drift correction and 80% with drift correction of all metabolites testing not significant (p_BH_ > 0.05).

For iPCA missing values were imputed with *missMDA::imputePCA(., ncp = 10)*^25^ and analysis was performed scaled and centred to unit variance (z-scaled) with *mixOmics::ipca(., scale = TRUE, ncomp = 2, mode = “deflation”)*^26^.

For OPLS-DA missing values were imputed with *MetaboAnalystR::ImputeMissingVar(., method = “knn_var”)*^27^, data was scaled and centred to unit variance (z-scaled) with *MetaboAnalystR::Normalization(…, “AutoNorm”)* and models were calculated with *MetaboAnalystR::OPLSR.Anal(., reg = TRUE)* with a standard 7-fold cross-validation for the factor *disease*. Model stability was additionally verified with 1000 random label permutations by *MetaboAnalystR::OPLSDA.Permut(., num = 1000)*.

Pearson correlation were calculated for each metabolite (drift corrected, *log_10_*-transformed data) against each numeric clinical parameter (untransformed) with *correlation::correlation()*^28^. Results were filtered to retain only metabolites and clinical parameters with at least one significant correlation (p_BH_ < 0.05). Retained correlations were clustered by Lance-Williams dissimilarity update with complete linkage using *stats::dist()* and *stats::hclust().* Dendogram were sorted with *dendsort::dendsort()*^29^ at every merging point according to the average distance of subtrees and plotted at the corresponding heat maps of Pearson R with *pheatmap::pheatmap()*^30^.

For univariate analysis of significant changes within each metabolite for the factor *disease* (i.e. PH vs. HC/DC) generalized least squares models were fitted with *nlme::gls()*^31,32^ without confounders and with potential confounders (age, sex, BMI) by maximum likelihood. For analysis within each cohort *log_10_*-transformed data was used, for joint analysis over all cohorts drift corrected, *log_10_*-transformed data was used, thus constituting a nonlinear approach. The three most common potential confounders (age, sex, BMI) were added stepwise in all possible combinations and model performances were compared within each metabolite by lower AIC (Akaike information criterion; relative estimate of information loss), higher log-likelihood (goodness of fit), significance in log-likelihood ratio test comparing two models, quality of Q-Q plots, randomness in residual and direct comparison of t-ratios. All models with any confounder combination showed significant influence (p < 0.01) on selected few metabolites (13–39). The model with age + sex impacted most metabolites. However, a direct comparison of t-ratio revealed a very small impact of age or sex correction on results, and according to model parsimony models without confounders were reported throughout.

FFA/lipid-ratios were calculated with FFAs in the numerator and lipids in the denominator. The numerators were all possible, summed (not weighed) combinations of up to six FFA from 11 FFAs (most significant in univariate analysis PH versus HC/DC and best analytical quality): FFA C15:0, FFA C16:2, FFA C16:1, FFA C17:1, FFA C17:0, FFA C18:3, FFA C18:2, FFA C18:1, FFA C19:1, FFA C20:5 and FFA C20:1. The denominators were all possible, summed (not weighed) combinations of up to four lipids from eight lipids (no significant change in univariate analysis and best analytical quality): LPC 18:2, LPC 18:1, PC 36:4, PC 38:6, SM 34:2, SM 36:2, LPE 16:0, and LPE 18:1. The combination of all possible summed FFA numerator and summed lipid denominator yielded a total of 240 570 different FFA/lipid-ratios. All used FFA and lipids had no missing values. The technical variability RSD in QC was calculated for each FFA/lipid-ratio following the rules of error propagation from the single metabolites RSD of QC in drift corrected, *log_10_*-transformed data. All FFA/lipid-ratios were calculated once without and once with drift correction (both *log_10_*-transformed). Diagnostic performance was tested by ROC analysis with *pROC::roc(…, algorithm = 2)*^33^ based on our training cohort. Test data like in machine learning approaches was not needed here because ratios are directly calculated without any model training. Therefore logistic regression was performed fitting training data with *stats::glm(…, family = “binomial”).* The performance was evaluated on the validation cohort using *pROC::roc()*. The optimal threshold was determined with *pROC::coords(…, ,best.method = “closest.topleft”)* to determine sensitivity and specificity.

Survival analysis used either times since sampling or times since diagnosis defining confirmed death as endpoint while censoring all others at time of last known follow-up. Impact of all relevant clinical parameters (untransformed), all single metabolites (drift corrected, *log_10_*-transformed data) and best performing FFA/lipid-ratios (*log_10_*-transformed data without drift correction) was analysed. FPHR4p scores were inverted so that higher values represent higher mortality risk. Numerical parameters were split into *high* and *low* with maxstat as optimal cut-off for survival prediction as determined by *survminer::surv_cutpoint(…, minprop = 0.3)*^34^. Kaplan**–**Meier curves were fitted for each category with *survival::survfit()*^35^, differences were tested with *survminer::ggsurvplot()* and plots with time since diagnosis were truncated at 15 years for better comparability with times since baseline (i.e. time since sampling). The Cox HR analysis^36^ was calculated with *survminer::coxph()* for the confounders (age, sex, BMI), the COMPERA 2.0 score, FPHR4p, and RATIO1 alone or in combinations. Numeric factors were categorized into *high* and *low* same as for Kaplan-Meier curves. The combination of RATIO1 with the FPHR4p or COMPERA 2.0 score was done additively with the same weighting on scaled values from 0 to 1, rescaling after addition.

Data visualisation and calculation of the machine learning and Cox HR analysis was performed with Python 3.9 (using the packages pandas, numpy, seaborn, sklearn, matplotlib, xgboost)^37–40^.

For machine learning, the package sklearn^41^ was used for the random forest (RF) and package xgboost for the XGBoost^42^ implementation, for better reproducibility a fixed random seed was set. Data was normalized with mean 0 and variance 1. A hyperparameter search for *number of trees {101, 301, 1001, 2001, **3001**}* and *depth {5, 10, 100, 200**, 300**}* for RF and *eta {0.1, **0.01**, 0.001}*, *depth {5, **10**, 100, 200, 300}* and *n_estimators {101, 301, 1001, 2001, **3001**}* for XGBoost was conducted with finally used hyperparameters highlighted in bold. Models were trained on training cohort data, which was randomly further divided five times into 80% for training and 20% for testing (stratified by class, age, sex, with non-overlapping test data). Trained models were validated with the external validation cohort, which had no data overlap with the training cohort.

Additionally to the original split by center (i.e. city of sample origin), all samples were artificially split into 70% training and 30% validation sets with balanced distributions in age, BMI, sex and class (PH/DC/HC) to overcome the potential bias from the unequal distribution of age, BMI, sex and class in the original training and validation cohorts by center. The distribution of age and BMI was equal according to a t-test as well as for sex and class (PH/HC/DC) according to a χ^2^ test (p > 0.2). All machine learning and FFA/lipid-ratio ROC analysis were repeated for these 70:30 training and validation sets.

Language editing was aided by the artificial intelligence tool https://instatext.io/ (last accessed November 2023).

## Supplementary Figures

**Fig. S1.**
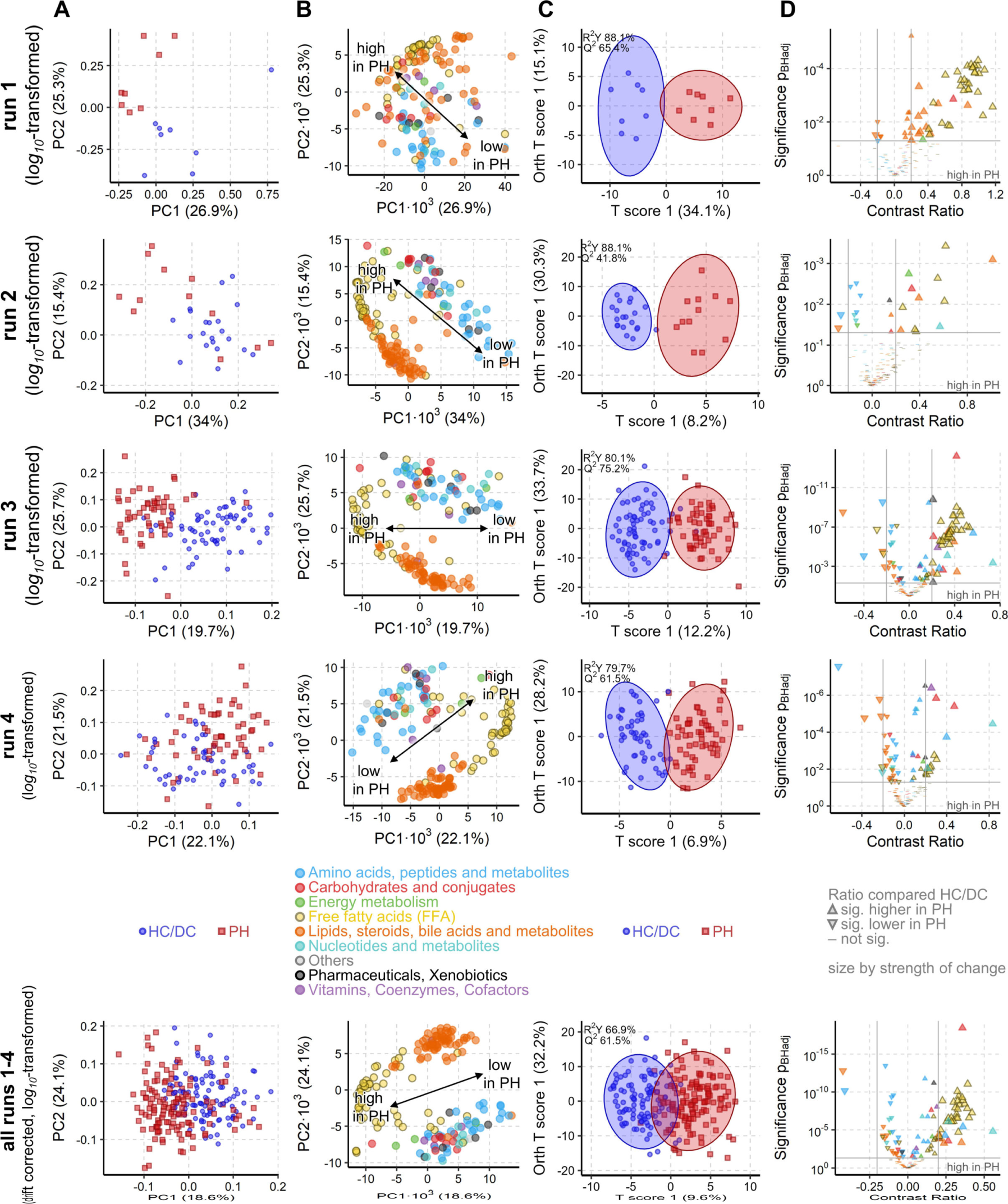
PH is associated with a strong metabolic shift in every measurement run. (A) iPCA scores plot representing the metabolic profile of each sample as a dot. The proximity of the dots indicates the similarity of the subjects’ metabolomes. Clear group separation by PH is visible along the first and or second component for each run and when all runs are jointly analyzed. (B) To A corresponding loadings plot in which each dot represents the contribution of the metabolite to the group separation observed in the scores plot. Free fatty acids (FFA, yellow circles) strongly drive the group separation and are increased in PH patients. (C) OPLS-DA maximizes the group difference from PH to HC/DC and the resulting scores plot represents, as in A, with dots the metabolome of each subjects. Similarly, proximity indicates similarity and ellipses mark the 95% confidence interval of the groups. The difference between the metabolome of PH and HC/DC was significant (Q^2^ > 50%, p< 0.001). (D) Volcano plot of univariate analysis highlighting significant (p_BH_ < 0.05, grey horizontal line) and strong (absolute contrast ratio > 0.25, grey vertical lines) increase in FFAs. For all methods 164 known metabolites and all samples from the measurement run per cohort (n_1_ = 16, n_2_ = 33, n_3_ = 118, n_4_ = 109) on *log_10_*-transformed was used. In run 4 measurement all HC samples from run 3 were repeated to evaluate reproducibility of samples across measurement runs.

**Fig. S2.**
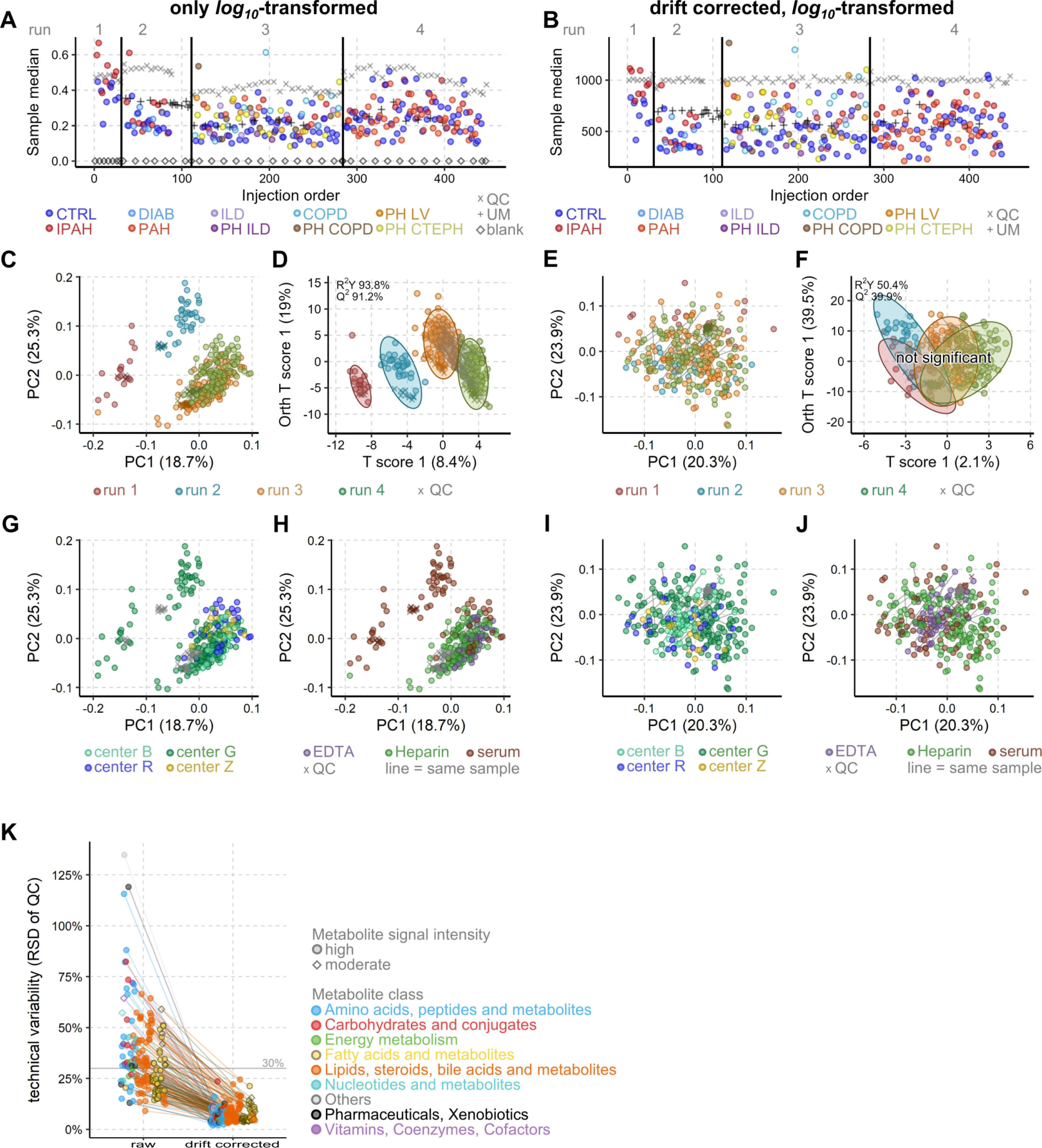
Drift correction improves data quality enabling a unified analysis of all 4 measurement runs. **(A, B)** Plot of the sample’s median signal intensity of all metabolites versus the sequence of measurement with measurements runs being separated by vertical black lines. **(A)** The QC samples (grey x) fluctuate within each measurement (= drift), while the batch effect is well visible as signal intensity jumps between the measurements. **(B)** Drift correction with an RF-based algorithm on QC samples removed drift and batch effects from the median sample intensity. **(C, E, G, H, I, J)** iPCA scores plots plot representing the metabolic profile of each sample as a dot. The proximity of the dots indicates the similarity of the metabolomes. Lines connect replicate measurements from same samples. Note **(C, G, H)** are the same iPCA model highlighting different biological factors. Analog **(E, I, J)** are the same iPCA model. **(C)** A clear group separation by measurement run is well visible between all 4 runs, with runs 3 and 4 being more similar due to the back to back measurement and shared HC and QC samples. **(D)** Drift correction removes the separation by measurement run. (D, F) OPLS-DA maximizes the differences between runs 1 to 4 and the resulting scores plot represents, as in C, with dots the metabolome of each sample. Similarly, proximity indicates similarity and ellipses mark the 95% confidence interval of the groups. **(D)** All 4 measurement runs were highly significantly (Q^2^ > 50%, p < 0.001) different before drift correction and **(F)** become non-significant after drift correction. **(G, I)** The observed group separation by sample origin (study center) before drift correction was significantly reduced after drift correction. **(H, J)** The observed group separation by sample material type before drift correction was significantly reduced after drift correction. **(K**) The strong reduction of the technical variability over all 4 runs after drift corrections shows how well the RF-based algorithm reduced technical noise. All plots are based on 164 known metabolites and all samples from the measurement run per cohort (n_1_ = 16, n_2_ = 33, n_3_ = 118, n_4_ = 109). A, C, D, G, H used *log_10_*-transformed. B, E, F, I, J used *log_10_*-transformed, drift corrected data.

**Fig. S3.**
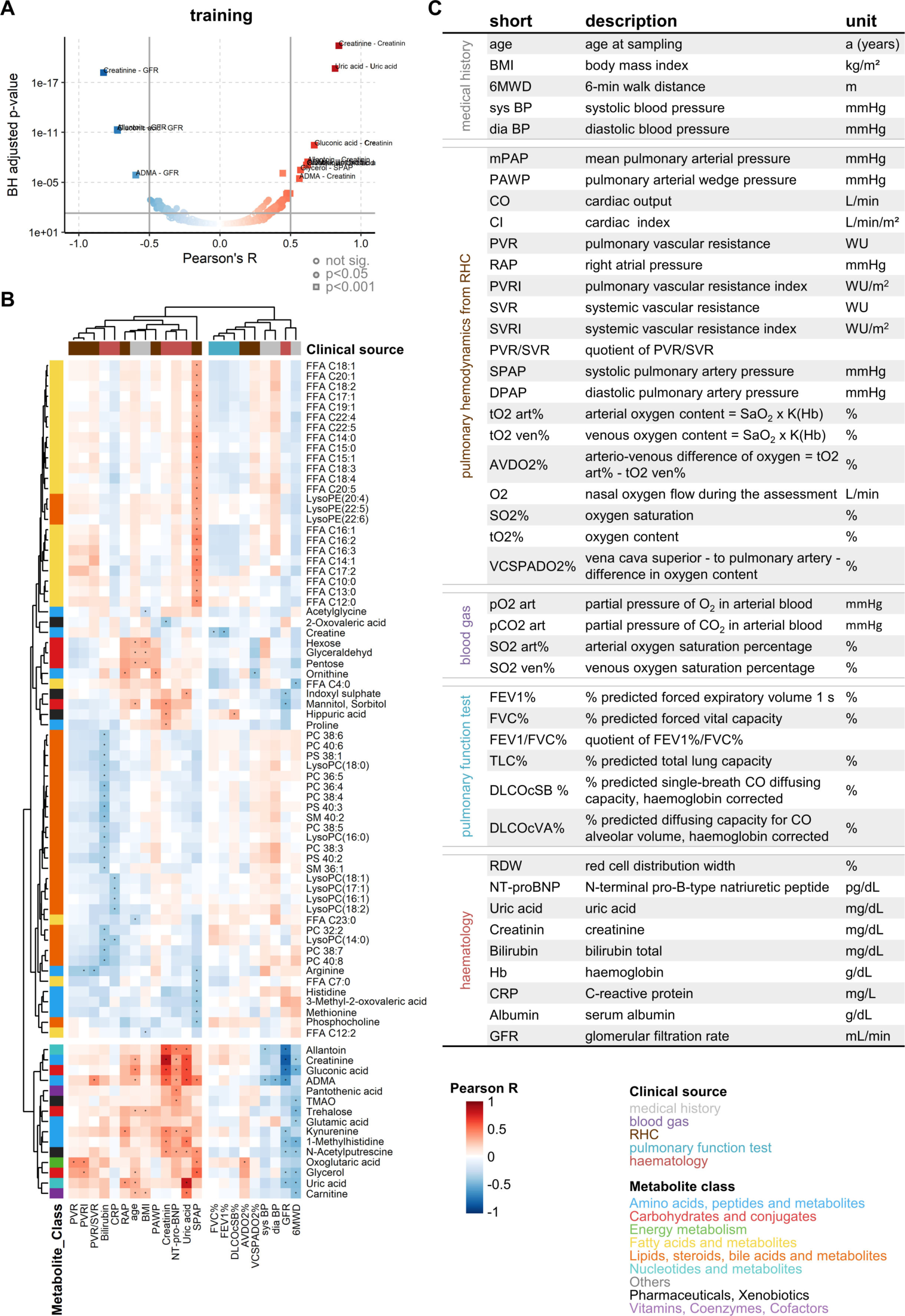
Correlation of selected metabolites with clinical parameters in the training cohort. **(A)** Volcano plot of all pairwise Pearson correlations of all 164 metabolites (drift corrected, *log_10_*-transformed data) versus 43 clinical parameters highlighting strong (absolute R > 0.5, grey vertical lines) and significant (p_BH_ < 0.05, grey horizontal line) correlations. Based on training cohort (n = 169). **(B)** Heatmaps with hierarchical clustering of the respective metabolite vs. clinical parameter. Pearson correlations were filtered to keep only rows and columns with at least one sig. correlation. All pairwise Pearson correlation results can be found in Supplementary Data 1. Uric acid and creatinine were measured as metabolite and were part of routine hematology. Accordingly, these parameters show the strongest, most significant correlations. **(C)** Overview of all investigated numeric clinical parameters correlated in (A, B) with all metabolites, listing their short names, explanations and units.

**Fig. S4.**
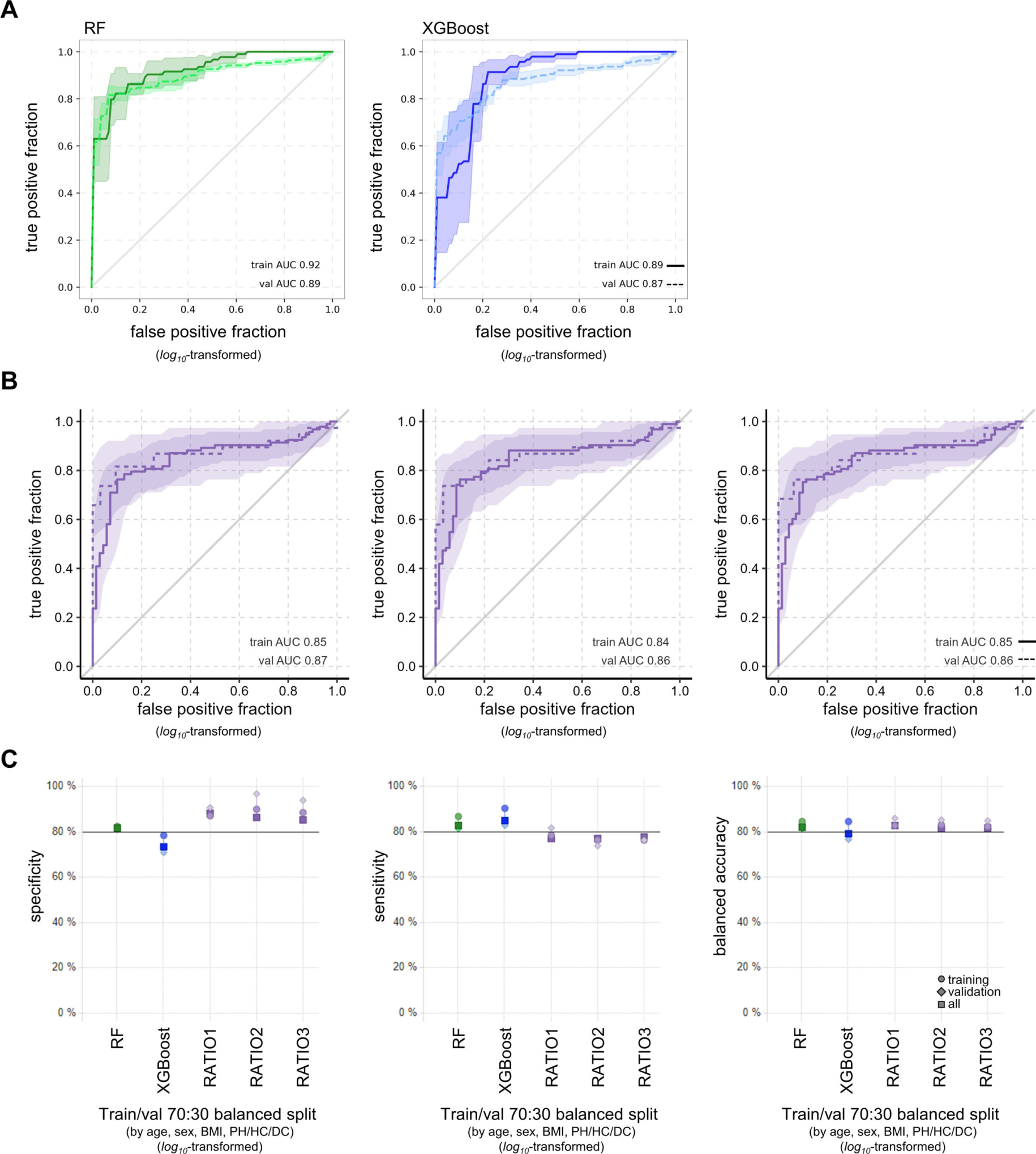
Data split has little impact on performance of RF, XGBoost or Ratios in predicting PH. The data was split 70:30 into trainings (n = 163, solid line) and validation set (n = 70, dashed line) balanced by age, BMI, sex and class, data was log_10_-transformed. **(A)** ROC plots of RF (green) and XGBoost (blue) trained with data from training cohort predicting class in validation cohort based on 153 metabolites and 95% confidence intervals marked by ribbons. **(B)** The ROC plots of the three best FFA/lipid-ratios and 95% confidence intervals marked by ribbons. **(C)** Plot of model performance metrics specificity, sensitivity and balanced accuracy for RF, XGBoost, and the three best FFA/lipid-ratios when based on either training (circles) or validation (diamonds) cohorts only or all available data (squares).

**Fig. S5.**
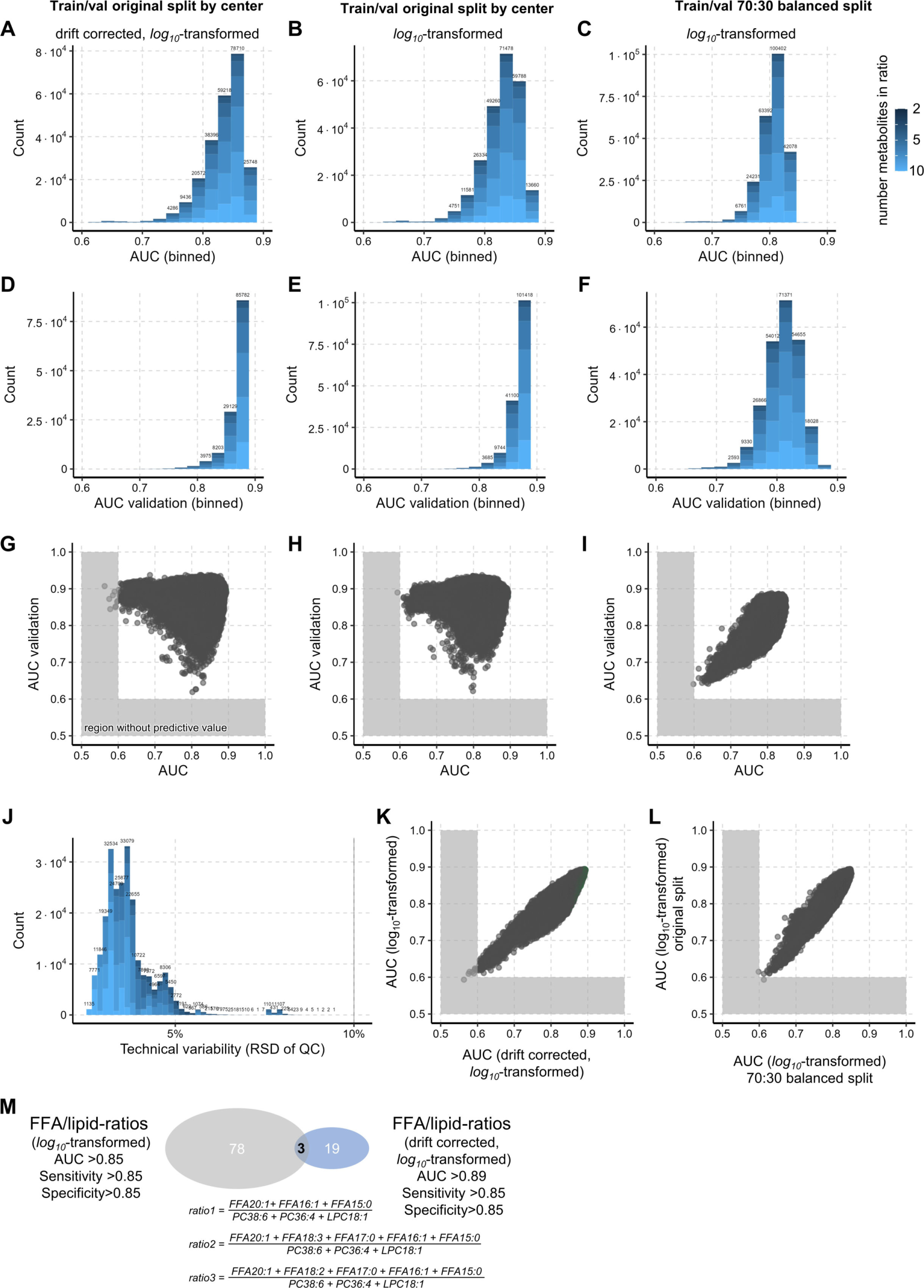
A quarter million of FFA/lipids-ratios could diagnose PH. **(A-F)** The histograms summarize the AUCs of the ROC analysis for all 240 570 FFA/lipid-ratios in each data type and split. The histogram of ROC analysis performance AUC for all possible FFA/lipid-ratios exhibits a tight distribution showing that all ratios performed similarly good. **(G-I)** Direct comparison of AUCs between the trainings and validation cohorts. **(A, B, G, H)** ROC analysis within the training cohort (cohort 1, 2, 3 n = 169) of the original split by centers benchmarked against the **(D, E, G, H)** validation cohort (cohort 4 n = 64). **(C, I)** ROC analysis within the trainings set (n = 163) of the 70:30 split (balanced by age, BMI, sex and class PH/DC/HC) and the **(F, I)** validation set (n = 70). **(A, D, G)** are based on drift *log_10_*-transformed, drift corrected data while **(B, C, E, F, H, I)** are based on *log_10_*-transformed data. **(J)** The histogram of the calculated technical error ratio for each ratio exhibits a tight distribution around 5% total technical relative variability showing. The grey vertical line marks the maximally acceptable technical variability cut-off for measurement methods in clinical routine laboratories. **(K)** The AUC from **(A)** and **(B)** are very similar to each other, confirming that the drift correction only slightly influences ROC analysis. **(L)** The AUC from **(B)** and **(C)** are very similar to each other, confirming that the split by center delivers very similar results to the 70:30 balanced split. Axes were scaled to exclude empty regions in **(A-I, K, L)**, especially for AUC < 0.6 which have no to very little predictive value. **(M)** Overview of top performing ratios for the original split by center with and without drift correction. The equations for the in both datasets top 3 ratios are given.

**Fig. S6.**
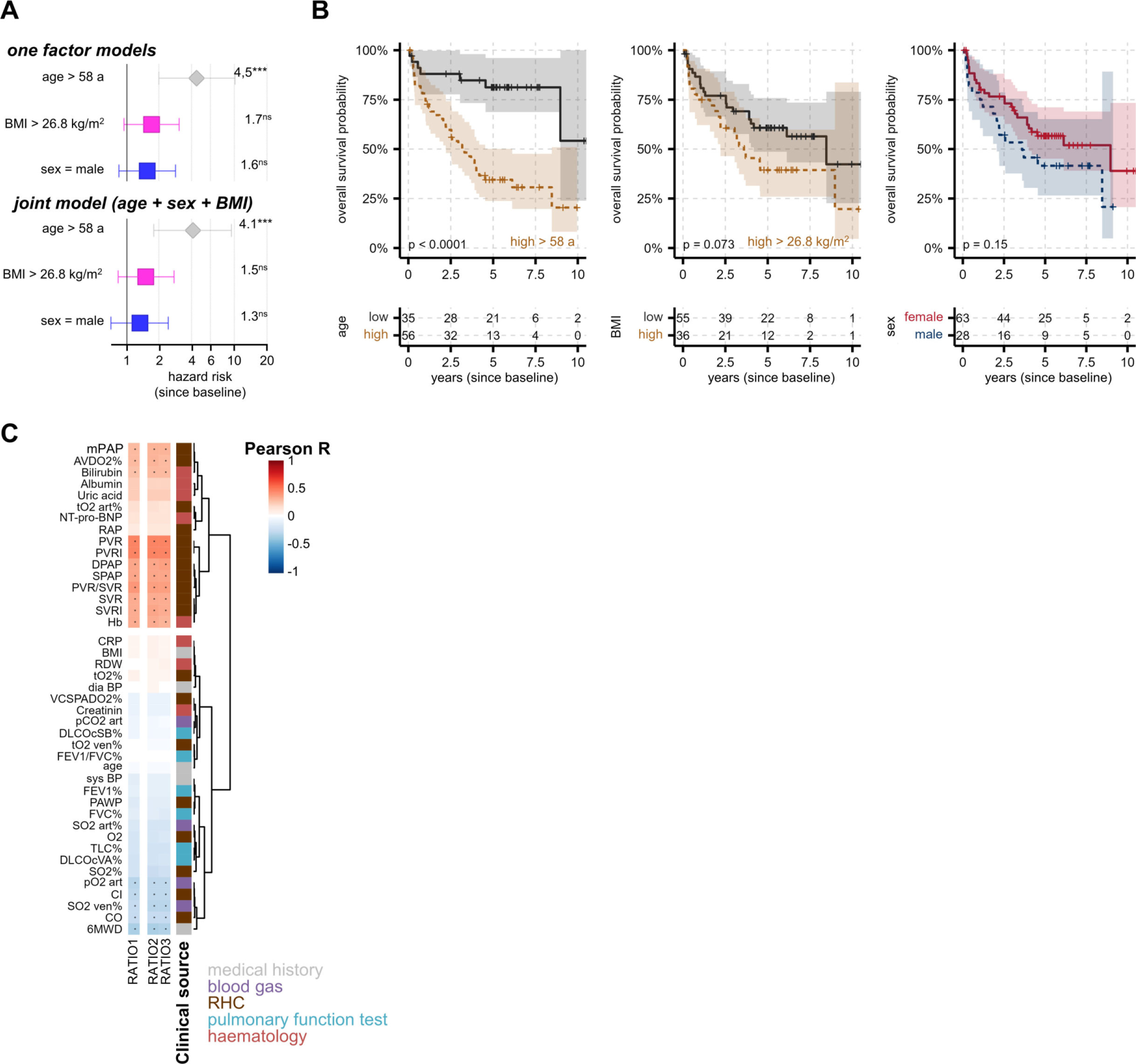
Survival analysis of covariates age, sex and BMI and linear correlation of top three RATIOs with clinical parameters. **(A)** Cox HR analysis for survival since baseline and the 95% confidence interval with statistical significance coded as not significant (ns); * p < 0.05; ** p < 0.01; *** p < 0.001. Higher age alone and in the tri-factor model was a significant risk factor while BMI and sex were slightly elevated but never significant. **(B)** Kaplan–Meier curves of survival times since baseline by age, sex or BMI. Age and BMI cut-offs were optimized with maxstat. **(C)** Heatmaps with hierarchical clustering of pairwise Pearson correlations of the top three RATIOs (*log_10_*-transformed data) versus 43 clinical parameters. All PH patients with survival times and both clinical scores were included (n = 91). All pairwise Pearson correlation results can be found in Supplementary Data 1.

**Fig. S7.**
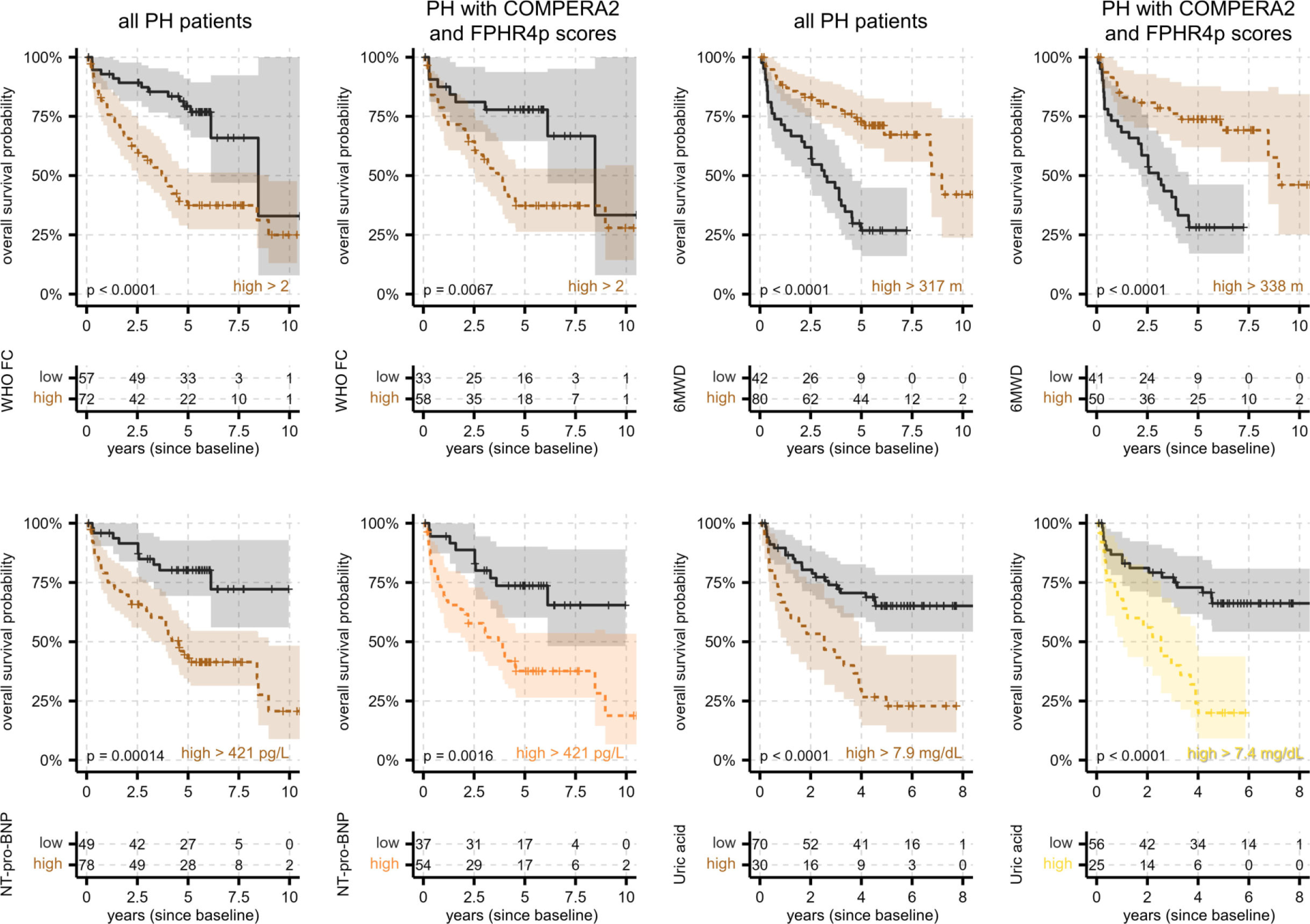
Survival analysis of established clinical risk factors and top metabolites. Kaplan–Meier curves analyzing survival times since baseline either for all PH patients or PH patients with both clinical scores available (n = 91). The stated cut-off for survival prediction was optimized with maxstat. **(A)** Survival of PH patients significantly decreases with known clinical risk factors such as higher WHO FC classes, lower 6MWD, higher NT-pro-BNP levels, and higher uric acid levels.

## Supplementary Tables

**Table S1:** Subject characteristics with medians ± 95% confidence intervals within measurement runs of the training cohort.

|  | run 1 |  | run 2 |  |  | run 3 |  |  |
| --- | --- | --- | --- | --- | --- | --- | --- | --- |
|  | HC (n=8) | PH (n=8) | HC (n=12) | DC (n=9) | PH (n=12) | HC (n=45) | DC (n=21) | PH (n=52) |
| Age at sampling, y | 57.0±8.8 | 58.5±9.0 | 70.5±10.8 | 56.0±10.1 | 63.0±9.3 | 58.0±2.6 | 60.0±5.7 | 66.5±3.7 |
| Female:male (ratio) | 7:1 (7:1) | 7:1 (7:1) | 11:1 (11:1) | 7:2 (3.5:1) | 11:1 (11:1) | 26:19 (1.4:1) | 13:8 (1.6:1) | 30:22 (1.4:1) |
| BMI kg/m<sup>2</sup> | 22.6±2.7 | 24.8±3.4 | 25.8±3.0 | 30.7±5.5 | 25.5±3.5 | 24.1±1.0 | 22.5±3.2 | 25.7±1.3 |
| Diagnosis since y | - | 3.0±5.9 | - | - | 6.0±4.0 | - | 8.0±2.1 | 0.0±1.1 |
| *Pulmonary hemodynamics from RHC* |  |  |  |  |  |  |  |  |
| mPAP (mmHg)<br>mean pulmonary arterial pressure | - | 45.0±7.7 | - | - | 41.5±5.8 | - | 22.0±0.8<sup>#</sup> | 41.0±3.1 |
| PAWP (mmHg)<br>pulmonary arterial wedge pressure | - | 10.0±4.2 | - | - | 5.5±1.7 | - | 9.5±0.3<sup>#</sup> | 10.0±1.6 |
| CO (L/min)<br>cardiac output | - | 4.1±1.6 | - | - | 4.3±0.8 | - | 3.9±0.4<sup>#</sup> | 4.6±0.5 |
| CI (L/min/m<sup>2</sup>)<br>cardiac index | - | 2.3±0.7 | - | - | 2.5±0.5 | - | 2.4±0.3<sup>#</sup> | 2.6±0.2 |
| PVR (WU)<br>pulmonary vascular resistance | - | 8.3±3.7 | - | - | 8.9±3.1 | - | 3.2±0.3<sup>#</sup> | 6.0±1.2 |
| RAP (mmHg)<br>right arterial pressure | - | 5.0±3.8 | - | - | 5.0±2.4 | - | 5.0±1.1<sup>#</sup> | 7.0±1.5 |
| *Clinical data* |  |  |  |  |  |  |  |  |
| 6MWD (m)<br>6-min walk distance | - | 338±96 | - | - | 431±149 | - | 454±50 | 317±32 |
| WHO FC<br>world health organisation functional class | - | 3.0±0.6 | - | - | 3.0±0.5 | - | 2.0±0.3 | 3.0±0.2 |
| FEV1 (% predicted)<br>forced expiration 1 s | - | 83.0±16.5 | - | - | 82.5±12.9 | - | 51.5±12 | 68.5±6.7 |
| FVC (% predicted)<br>forced vital capacity | - | 90.9±18.0 | - | - | 97.6±14.8 | - | 63.8±8.2 | 78.0±6.5 |
| FEV1/FVC (% predicted) | - | 73.3±6.0 | - | - | 78.3±6.5 | - | 56.3±10.3 | 73.2±3.9 |
| TLC (% predicted)<br>total lung capacity | - | 95.7±12.5 | - | - | 103.1±9.4 | - | 103.0±13.2 | 92.0±5.2 |
| DLCO cSB (% predicted)<br>single-breath CO diffusing capacity,<br>hemoglobin corrected | - | 69.5±8.4 | - | - | 65.4±9.5 | - | 49.6±9.6 | 56.4±8.2 |
| DLCO cVA (% predicted)<br>CO diffusing capacity alveolar volume,<br>hemoglobin corrected | - | 74.6±14.1 | - | - | 60.6±20.8 | - | 70.0±9.6 | 71.0±7.8 |
| RDW (%)<br>red cell distribution width | - | 15.5±1.1 | - | - | 14.4±0.9 | - | 14.0±0.9 | 15.7±1.0 |
| NT-proBNP (pg/mL) | - | 508±1197 | - | - | 263±1122 | - | 98±47 | 1100±1112.2 |
| Uric acid (mg/dL) | - | 5.7±1.4 | - | - | 5.7±1.6 | - | 5.0±0.6 | 6.2±0.7 |
| Creatinine (mg/dL) | - | 0.92±0.17 | - | - | 1.02±0.19 | - | 0.80±0.11 | 1.04±0.17 |
| Bilirubin total (mg/dL) | - | 0.61±0.25 | - | - | 0.57±0.26 | - | 0.40±0.16 | 0.70±0.17 |

<sup>#</sup>based on 4 patients, not representative

**Table S2:** Characteristics of patients with IPAH and healthy donors used for laser capture-microdissection of PA.

|  | ID | Age | Sex |  | ID | Age | Sex | mPAP |
| --- | --- | --- | --- | --- | --- | --- | --- | --- |
| Donor | 1 | [50-59] | male | IPAH | 1 | [30-39] | female | 50 |
|  | 2 | [70-79] | female |  | 2 | [30-39] | female | 88 |
|  | 3 | [50-59] | female |  | 3 | [18-29] | female | 56 |
|  | 4 | [30-39] | male |  | 4 | [18-29] | female | 69 |
|  | 5 | [18-29] | female |  | 5 | [50-59] | male | 66 |
|  | 6 | [50-59] | male |  | 6 | [18-29] | male | 74 |
|  | 7 | [18-29] | male |  | 7 | [50-59] | male | 65 |
|  | 8 | [70-79] | female |  | 8 | [30-39] | female | 69 |
|  | 9 | [50-59] | female |  | 9 | [40-49] | male | 96 |
|  | 10 | [50-59] | female |  | 10 | [18-29] | female | 65 |

**Table S3:**
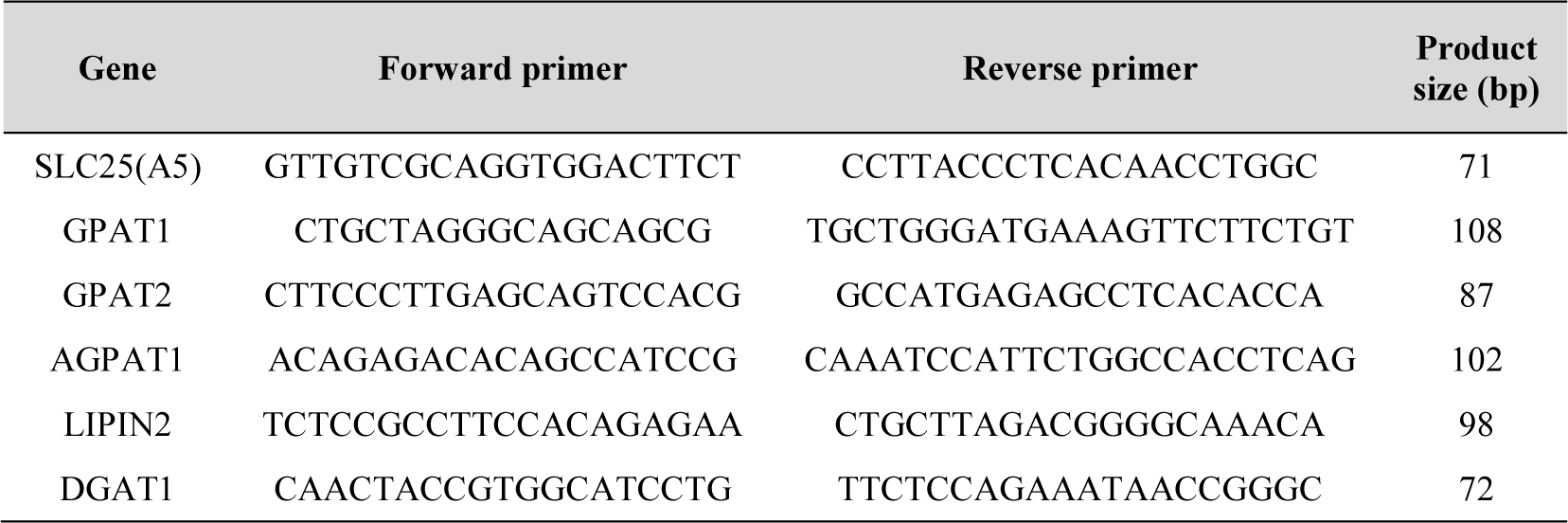
Primers used to assess the expression of listed genes. Gene name, PubMed Nucleotide accession number used for primer design, forward and reverse primer sequences and the size of the PCR product (in bp) 4 are given. All primers were designed so that the PCR product span at least one exon-exon junction.

**Table S4:** Characteristics of the hPASMC and hPAEC donors used for in vitro studies.

|  | ID | Age | Sex |  | ID | Age | Sex |
| --- | --- | --- | --- | --- | --- | --- | --- |
| hPASM | 1 | [30-39] | male | hPAEC | 1 | [50-59] | male |
|  | 2 | [50-59] | female |  | 2 | [40-49] | female |
|  | 3 | [30-39] | male |  | 3 | [50-59] | female |
|  | 4 | [30-39] | female |  | 4 | [50-59] | male |
|  | 5 | [18-29] | female |  |  |  |  |
|  | 6 | [30-39] | male |  |  |  |  |

